# Functional alterations of two salience-related systems jointly and independently contribute to psychosis

**DOI:** 10.1101/2021.10.02.21264326

**Authors:** Jun Miyata, Toby Winton-Brown, Thomas Sedlak, Toshihiko Aso, Nicola Cascella, Jennifer Coughlin, Nicolas A. Crossley, Emrah Duezel, Takahiro Ezaki, Masaki Fukunaga, Carolyn Howell, Masanori Isobe, Kouhei Kamiya, Kiyoto Kasai, Takanori Kochiyama, Shinsuke Koike, Akira Kunimatsu, Naoki Masuda, Susumu Mori, Yasuo Mori, Toshiya Murai, Kiyotaka Nemoto, Frederick Nucifora, Kazutaka Ohi, Naohiro Okada, Yuki Sakai, Nobukatsu Sawamoto, Tsutomu Takahashi, Shinichi Urayama, Yoshiyuki Watanabe, Crystal C. Watkins, Hidenaga Yamamori, Yuka Yasuda, Ryota Hashimoto, Hidehiko Takahashi, Akira Sawa, Philip McGuire

## Abstract

**Background and hypothesis:** Salience is a critical mechanism for animal survival. Aberrant salience is essentially implicated in psychosis, involving the midbrain–striatum–hippocampus and the anterior cingulate cortex (ACC)–insula salience network (SN) systems. However, whether these two systems jointly or independently contribute to psychosis traits, staging, and state remains unknown.

**Study design:** Eight scanners at seven institutions recruited 209 subjects with psychosis at the ultra-high-risk (UHR), first-episode (FEP), and chronic (ChrP) stages and 279 matched healthy controls (HC). Resting-state functional MRI data, which were intensively denoised and scanner effect-removed, revealed the two systems comprising five networks: midbrain-thalamic and striatal parts of the basal ganglia network (BGN-MbThal and BGN-Str), the medial temporal lobe network (MTLN), and the ACC and insular parts of the SN (SN-ACC and SN-Ins). Group differences, correlations with positive symptoms, and effects of medication/psychosis type of the networks were investigated in each psychosis stage.

**Study results:** Connectivity within the SN-ACC was reduced in UHR compared to HC (p<0.05, family-wise-error [FWE] corrected) and in the FEP and ChrP stages at liberal thresholds, with effect sizes of UHR>FEP>ChrP. FEP showed reduced connectivity within the BGN-MbThal, increased brain-state instability among the five networks, and positive correlations between positive symptoms and connectivity within and between the MTLN (all p<0.05, FWE). The correlation was stronger in unmedicated than in medicated, and in affective than in non-affective psychosis patients (p<0.05, FWE).

**Conclusions:** The results provide a novel concept that the two salience mechanisms work in an integrated and separate manner in psychosis.

## Introduction

Salience denotes being particularly noticeable or important, and is computationally defined as the absolute value of the prediction error^1^ and the Laplacian of stimuli^2^. For survival, animals must quickly allocate their limited attentional resources to salient stimuli in the surrounding world. Previous animal studies showed that midbrain-striatal dopaminergic activity codes motivational (reward and avoidance) salience^3–5^, whereas human functional magnetic resonance imaging (fMRI) studies showed that the striatum is responsive to stimulus salience^6–9^.

Alterations in the midbrain-striatum region are indicated in psychosis^10^, most commonly schizophrenia^11^. Elevated dopamine levels in the striatum of patients with schizophrenia^12^ have been hypothesized to cause aberrantly heightened salience attribution to ordinary stimuli, leading to delusions and hallucinations; i.e. positive symptoms^13^ (aberrant salience hypothesis)^10^. Additionally, recent circuit models of psychosis indicate that hippocampal overactivity triggers a hyperdopaminergic state in the midbrain and striatum^14^. These findings were consistent with those of recent clinical studies^15,16^. Thus, the midbrain-striatum-hippocampus system plays a critical role in aberrant salience in psychosis.

Human fMRI studies have described another neural system, the salience network (SN), which mainly comprises the anterior cingulate cortex (ACC) and bilateral insula and is activated by the salience of stimuli rather than the nature of tasks^17,18^. Recent studies have implicated SN alterations in psychotic symptoms and schizophrenia^19–23^. However, a critical unanswered question is the role of the ACC-insular SN system in the aberrant salience of psychosis. Alternatively, do these two salience-related systems contribute to aberrant salience either jointly or differently?

Recently, the SN is included in the “triple network model”, which postulates that the SN act as a switching-hub between the default mode network (DMN) that is activated during rest/introspection, and the frontoparietal network (FPN) that is activated on task/cognitive effort^24,25^. Dysfunction of the triple network is associated with various psychiatric disorders, including psychosis^26–29^. However, elucidating the pathophysiology of aberrant salience in psychosis requires consideration of the two neural systems directly related to salience per se. To date, no study has done so.

Resting-state fMRI (rsfMRI) is widely used to investigate the functional connectivity of brain networks in patients with psychiatric disorders. Independent component analysis (ICA) of rsfMRI can identify large-scale networks representing the midbrain-striatum-hippocampus and ACC-insular SN systems in a data-driven manner; the midbrain and striatum were identified as the basal ganglia network (BGN^30^), the bilateral hippocampus was identified within the medial temporal lobe network (MTLN^31^, also known as meso/paralimbic network^32^), and the SN comprised the ACC and insula^17^. Investigating the connectivity within and between these three networks can answer the questions above.

Biomarkers for psychosis/schizophrenia are essentially needed. Biomarkers are generally categorized as 1) trait markers, which are present throughout the disease course and are not changed by treatment; 2) staging markers, which are prominent in certain disease stages; and 3) state markers, which reflect symptoms and changes according to the treatment^33–36^. The course of psychosis can be divided into ultra-high-risk (UHR), first-episode (FEP), and chronic (ChrP) psychosis stages^37^. Whether aberrant salience is associated with psychosis traits, stage, or state is unclear, although dopaminergic dysfunction is reportedly more marked in FEP than in UHR^38^.

Thus, we investigated how the two salience-related systems are associated with the aberrant salience of psychosis using ICA of rsfMRI. Our main outcome was whether functional connectivity within and between the BGN, MTLN, and SN was associated with the 1) diagnosis of psychosis and/or 2) positive symptom severity at the UHR, FEP, and ChrP stages. The secondary outcomes were whether antipsychotic medication and psychosis type (affective or non-affective psychoses) affected the findings of 1) and 2), to further clarify the findings. To fully describe the connectivity profile, we investigated two complementary aspects of static (temporally constant across the time-course)^39^ and dynamic^40^ (temporally varying during the time-course) network measures. Considering that marked dopaminergic dysfunction in FEP and the effects of antipsychotics (dopamine blockers) on positive symptoms, we hypothesized that the midbrain-striatum-hippocampus system would be associated with positive symptoms as well as medication, especially in FEP.

## Methods

### Participants

The details are described in the Supplemental Methods and Supplemental Tables S1−S7 in the Supplemental Material. We recruited 488 individuals with UHR, FEP, and ChrP and matched healthy controls (HC) from eight scanners at seven institutions (Table 1). Hereafter, we refer to individuals with UHR as “patients” to refer to UHR, FEP, and ChrP as a whole.

**Table 1.**
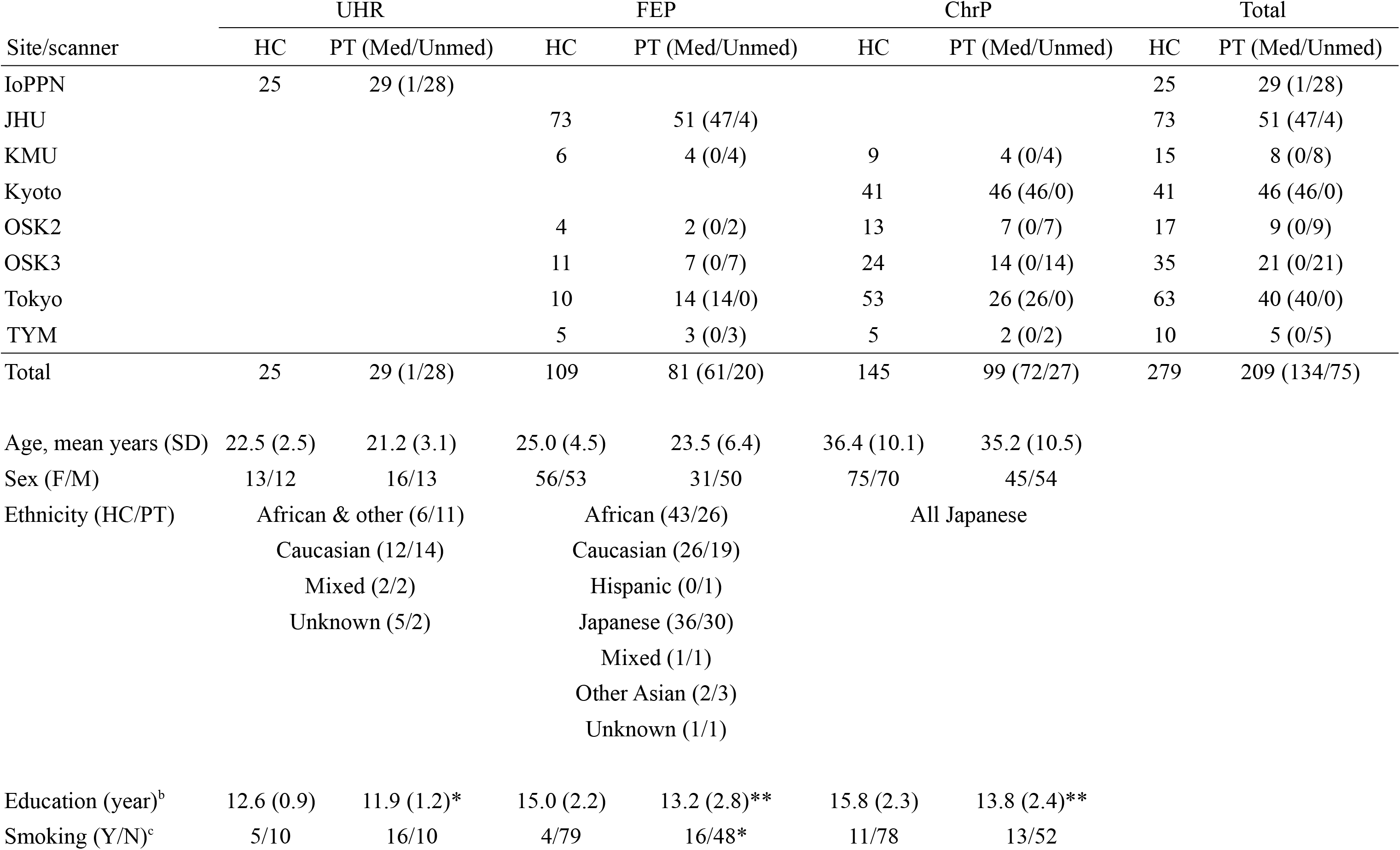
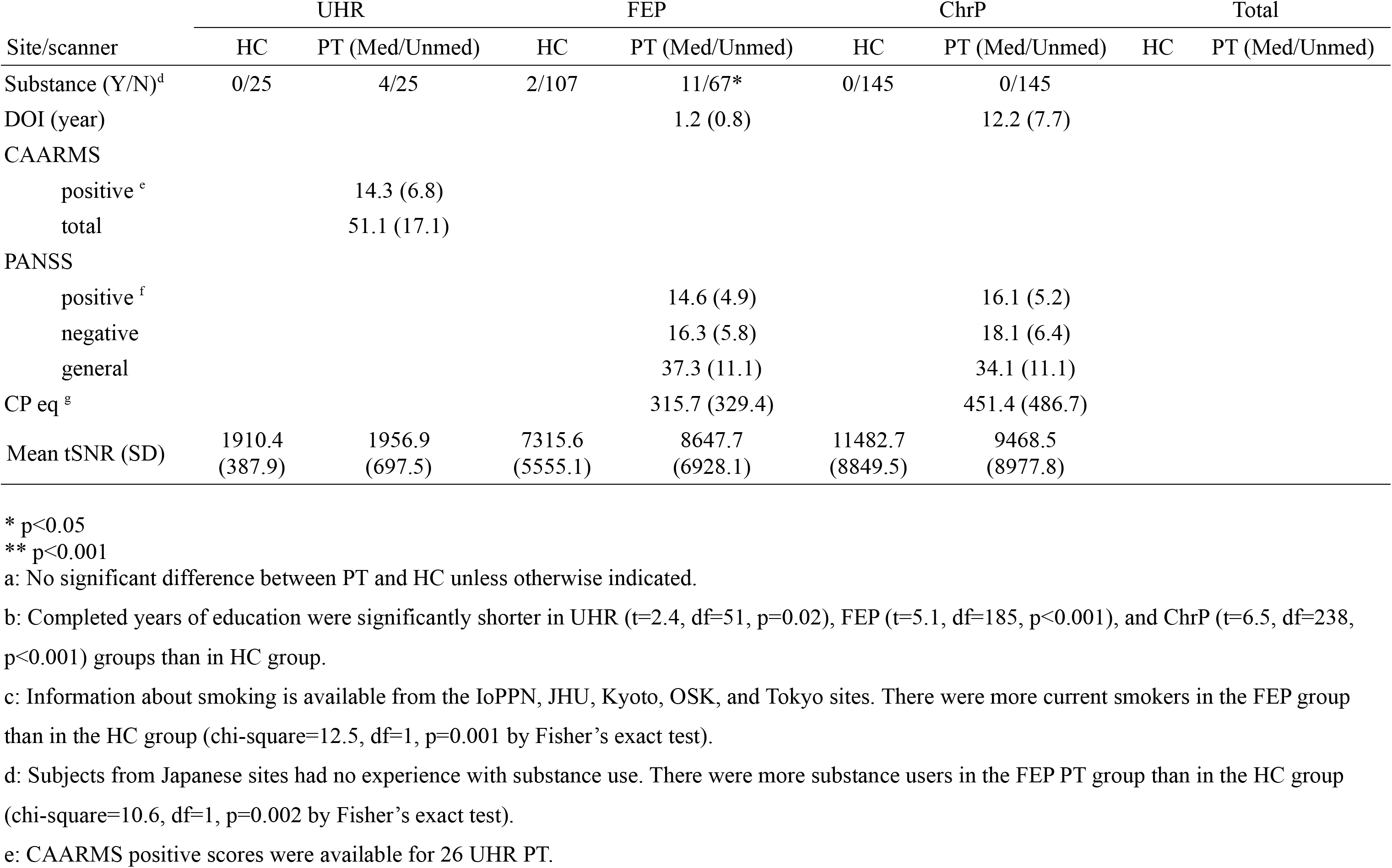
Participant demographic and clinical characteristics.^a^.

The medication status (unmedicated or medicated) and psychosis type (affective or non-affective) of FEP were partially unbalanced, while those of ChrP were unbalanced.

After receiving a complete description of the study, all participants provided written informed consent. The study design was approved by the following institutional review boards of each institution:

approved by Hammersmith Hospital Research Ethics Committee (IoPPN);

approved by Johns Hopkins Medicine Institutional Review Board (JHM IRB) (JHU);

approved by Research Ethical Committee of Kanazawa Medical University (KMU);

approved by Kyoto University Graduate School and Faculty of Medicine, Ethics Committee (Kyoto);

approval by Research Ethics Committee, Osaka University (OSK);

approved by Research Ethics Committee of the Faculty of Medicine of the University of Tokyo (Tokyo);

and approved by Ethics Committee, University of Toyama (TYM).

### MRI acquisition

The details are provided in Supplemental Methods and Supplemental Table S8.

### Preprocessing

The details are provided in the Supplemental Methods, Supplemental Table S9, Supplemental Figures S1 and S2, and Supplemental Video S1. In addition to conventional preprocessing steps, ICA-based denoising^41^ was performed to intensively remove non-neuronal noise from the rsfMRI data.

As an index of data quality, the temporal signal-to-noise ratio (tSNR) was calculated at each voxel as the mean/SD of the time course and was averaged for the whole brain using QAscript**^42^.**

### Meta-ICA to identify group-level networks

We performed meta-ICA^43,44^, which has higher robustness and reproducibility for group-level network identification than conventional single-shot group ICA^45^. The details are described in the Supplemental Methods.

### Network identification

After meta-ICA, the networks of interest (NOIs: the BGN, SN, and MTLN) were identified (Supplemental Methods, Supplemental Figure S3, and Supplemental Table S10). The BGN and SN were split into two parts, for a total of five NOIs (Figure 1a).

### Thresholded dual regression

Details are in provided in the Supplemental Methods. After meta-ICA, individualized spatial maps and time courses of each network were obtained using thresholded dual regression^46^ to more accurately estimate the time courses of networks than conventional dual regression^47^. Spatial maps of the five NOIs were used for static within-network connectivity analysis. For static between-network connectivity, Z-transformed partial correlations between the time courses of the NOI-pairs were calculated while controlling for the other three NOIs.

### Dynamic analysis by ELA

We performed dynamic^40^ analysis using energy landscape analysis (ELA)^48–53^ (Supplemental Methods), which estimated an “energy landscape” with two stable brain states from activity patterns of the time courses of the NOIs obtained above. States A and B were characterized by “all-NOIs-deactivated” and “all-NOIs-activated” patterns, respectively (Figure 1b and 1c). Then, two indices of the frequency and transition rate of the brain states were calculated for each participant.

**Figure 1.**
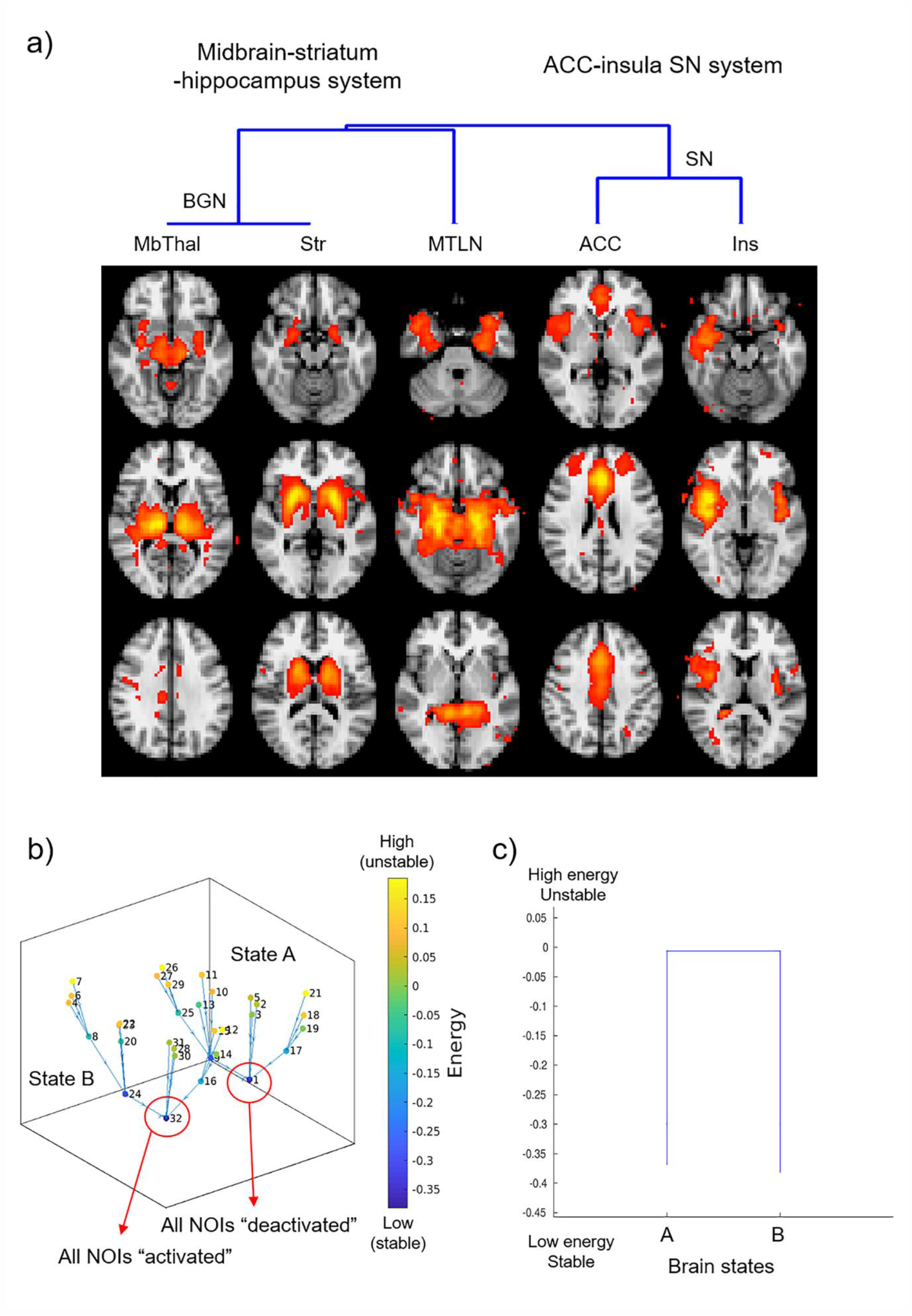
Networks of interest (NOIs) characteristics. a) Meta-ICA identified the midbrain-striatum-hippocampus system as the MbThal and Str parts of the BGN and the MTLN. The SN system was identified as the ACC and Ins parts. The hierarchical clustering is shown on top. b, c) Energy landscape analysis aggregated the time course patterns of the NOIs into two brain states. In the local minima (low-energy, frequent pattern) of state A, five NOIs were “deactivated” (below the mean time-course) in synchronization, but were “activated” (above the mean) in state B. **Abbreviations** ACC, anterior cingulate cortex; BGN, basal ganglia network; ICA, independent component analysis; Ins, insula; MbThal, midbrain and thalamus; MTLN, medial temporal lobe network; SN, salience network; Str, striatum.

### Site/scanner effect harmonization

Although multi-site, large sample-size studies can increase reproducibility^54^, site/scanner difference substantially affects their results^55–57^. Thus, we estimated and removed additive and multiplicative site/scanner effects using ComBat ^58–60^, which can also effectively harmonize unbalanced data^59^. The details are provided in the Supplemental Methods. Supplemental Figure S4 illustrates efficient removal of the site/scanner effect.

### Statistical analysis

Details are provided in the Supplemental Methods. Permutation-based non-parametric inference^61^ was used to perform 1) group comparisons between patients and HC and 2) multiple regression analyses with positive symptom severity, for within- and between-network connectivity and ELA indices at each psychosis stage. Age, sex, and mean tSNR were covariated. The statistical threshold was p < 0.05, family-wise error (FWE) corrected for the number of voxels, contrasts, and five networks/10 network pairs/two ELA indices. We also reported at more liberal thresholds (p < 0.1, FWE for voxels, contrasts, and networks/network pairs/indices and p < 0.05, FWE for voxels and contrasts).

### Effects of medication and psychosis type

We then investigated the effect of medication status and psychosis type on significant group differences and correlations, as described in the Supplemental Methods. In the case of FEP, since Johns Hopkins University (JHU) includes both medication status and psychosis types, we additionally performed the same analyses for this site only.

### Effects of comorbidity, smoking, and substance use

We investigated the effects of comorbidities, smoking, and substance use (Supplemental Methods).

## Results

### 1) Demographic data

The details of Table 1 are described in the Supplemental Results.

### 2) Main outcome 1: group comparisons

Compared to the HC, we observed significant connectivity reductions within the ACC part of the SN (SN-ACC) in UHR, and within the midbrain-thalamus part of the BGN (BGN-MbThal) in the right thalamus in FEP (p = 0.003 and 0.02, respectively; FWE corrected for voxels, contrasts, and networks) (Figure 2a). At more liberal thresholds, the SN-ACC showed a reduction in FEP (p = 0.07, FWE for voxels, contrasts, and networks) and ChrP (p = 0.04, FWE for voxels and contrasts) at almost the same locations as in UHR (Figure 2b). The effect sizes of these clusters for UHR, FEP, and ChrP were 1.45, 0.60, and 0.54, respectively (Figure 2c)^62^. Other results at liberal thresholds are shown in Supplemental Figure S5.

**Figure 2.**
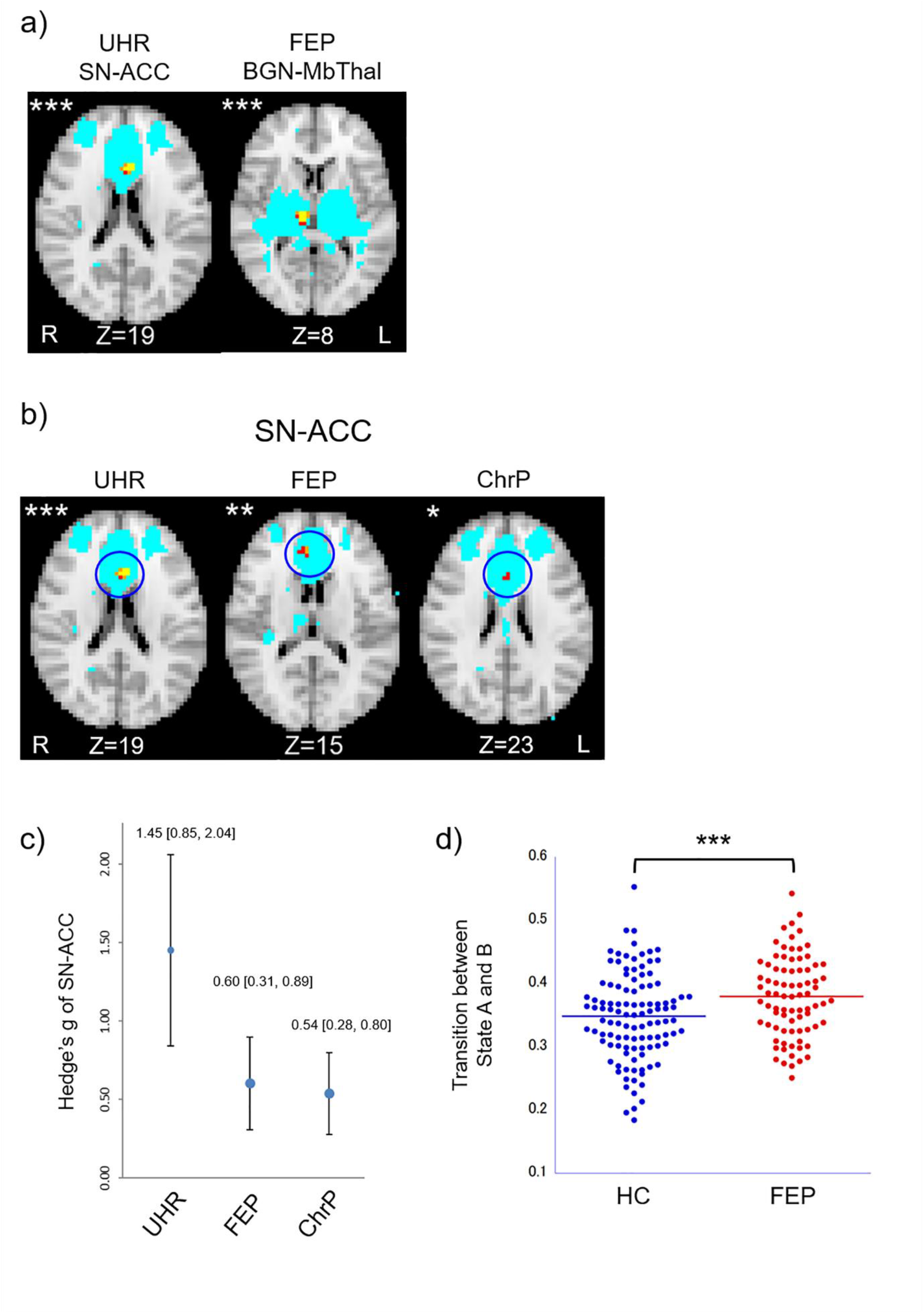
Group comparisons. a) Reduced within-network connectivity in UHR and FEP. b) The significance level of the SN was UHR > FEP > ChrP. Yellow/*** p < 0.05, family-wise error (FWE) corrected for voxels, contrasts, and networks. Orange/** p < 0.1, FWE for voxels, contrasts, and networks. Red/* p < 0.05, FWE for voxels and contrasts. Light blue: NOIs. c) The effect size of the ACC was UHR > FEP > ChrP. d) Increased transition between brain states in FEP. p < 0.05, FWE for contrasts and ELA indices. **Abbreviations** ACC, anterior cingulate cortex; BGN, basal ganglia network; ChrP, chronic schizophrenia; ELA, energy landscape analysis; FEP, first-episode psychosis; HC, healthy controls; Ins, insula; MbThal, midbrain and thalamus; MTLN, medial temporal lobe network; NOI, network of interest; PT, patients; SN, salience network; Str, striatum; UHR, ultra-high-risk for psychosis.

Between-network connectivity did not differ at any stage, even at liberal thresholds. ELA revealed a significantly increased transition between the two brain states in FEP (p = 0.003, FWE for contrasts and indices) (Figure 2d). No other significant differences were observed even at liberal thresholds.

### 3) Main outcome 2: correlational analyses

We observed a significant positive correlation between positive symptom severity (positive scale of Positive and Negative Syndrome Scale [PANSS]^63^) and connectivity within the MTLN in the left/right hippocampus and parahippocampal gyrus in FEP (p = 0.03 and p = 0.02 for the left and right, respectively; FWE for voxels, contrasts, and networks) (Figure 3a and 3b). Figure 3c shows no clear site/scanner effects. Supplemental Figure S6a shows the results at liberal thresholds. No other significant correlations were observed.

**Figure 3.**
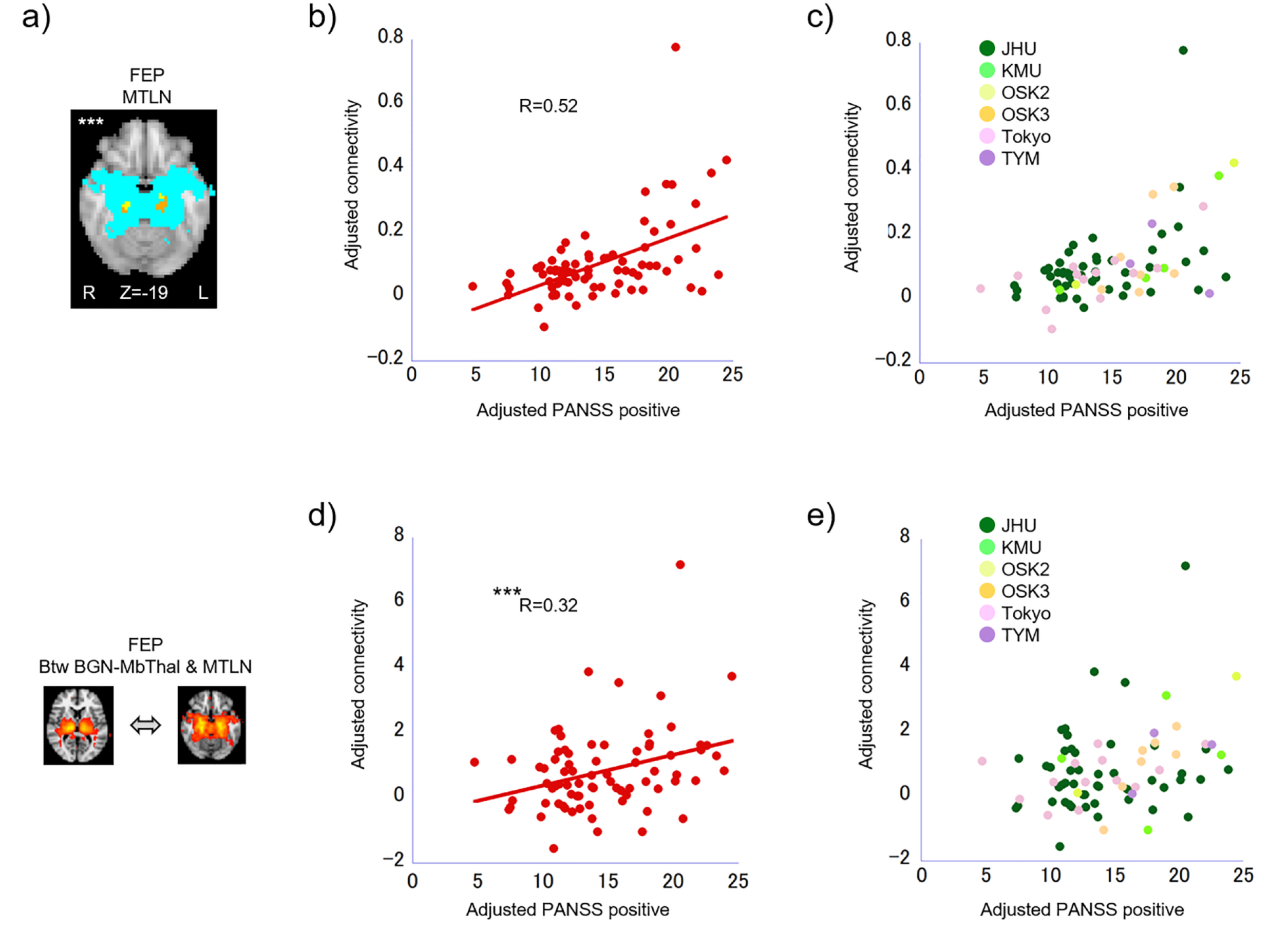
Correlational analysis results in FEP (n=76). a) Positive correlation between PANSS positive scale and the connectivity within the MTLN. Yellow/*** p < 0.05, family-wise error (FWE) corrected for voxels, contrasts, and networks. Orange: p < 0.1, FWE for voxels, contrasts and networks. Light blue: MTLN NOI. b) Scatter plot of a). c) No clear site/scanner effect. d) Positive correlation between the PANSS positive scale and connectivity between the MTLN and BGN-MbThal. *** p < 0.05, FWE for contrasts and network-pairs. e) No clear site/scanner effect. For b) – e), the PANSS and connectivity values were adjusted for age, sex, and tSNR. For the site/scanner names c) and e), please refer to Table 1. **Abbreviations** BGN, basal ganglia network; Btw, between; FEP, first-episode psychosis; MbThal, midbrain and thalamus; MTLN, medial temporal lobe network; NOI, network of interest; PANSS, Positive and Negative Syndrome Scale; tSNR, temporal signal-to-noise ratio.

A significant positive correlation was also found between the PANSS positive scale and connectivity between the MTLN and BGN-MbThal in FEP (p = 0.04, FWE for contrasts and network pairs) (Figure 3d). Figure 3e shows no clear site/scanner effects. Supplemental Figure S6b shows results at liberal thresholds for UHR and FEP. No other significant correlations were observed.

The ELA indices did not show significant correlations with positive symptoms even at liberal thresholds.

### 4) Secondary outcome 1: effect of medication

We did not find any significant effect of antipsychotic medication on group comparison results (Supplemental Results).

The regression coefficients between the PANSS and the mean cluster value of the MTLN were significantly larger in unmedicated patients than in medicated patients, both for all sites and JHU alone (p = 0.009 and p < 0.001, respectively, FWE for contrasts. Figure 4a top row).

**Figure 4.**
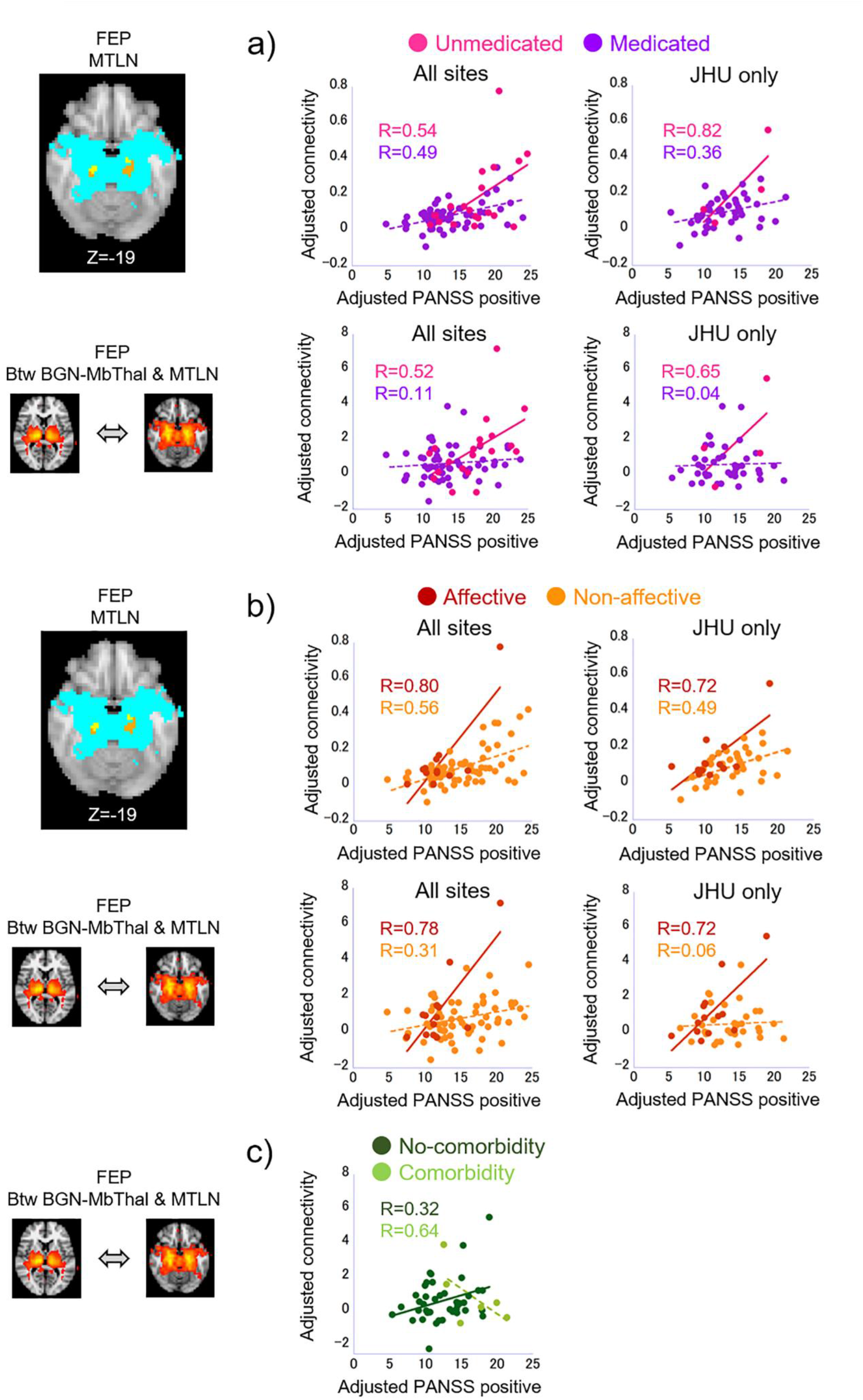
Effect of medication, psychosis type, and comorbidity. All p < 0.05, FWE for contrasts. The PANSS and connectivity values were adjusted for age, sex, and tSNR. a) Larger regression coefficients between the PANSS positive scale and MTLN connectivity in unmedicated patients than in medicated patients. Upper row: connectivity within the MTLN. Bottom row: connectivity between the MTLN and BGN-MbThal. Left column: all sites (unmedicated/medicated = 20/56). Right column: JHU only (unmedicated/medicated = 4/43). b) Larger regression coefficients between PANSS and MTLN connectivity in patients with affective psychosis compared to those with non-affective psychosis. The upper/lower rows and left/right columns are the same as in Figure 4a. Affective/non-affective psychosis = 13/63 and 13/34 for all sites and JHU only, respectively. c) Larger regression coefficient between PANSS and between-network connectivity of the MTLN in patients without (n = 41) than in those with (n = 6) comorbidity (JHU only). **Abbreviations** BGN-MbThal, midbrain and thalamus part of the basal ganglia network; Btw, between; FEP, first-episode psychosis; JHU, Johns Hopkins University; MTLN, medial temporal lobe network; PANSS, positive and negative syndrome scale; tSNR, temporal signal-to-noise ratio.

The regression coefficients between PANSS and the connectivity between the MTLN and BGN-MbThal were also significantly larger in unmedicated patients than in medicated patients for all sites and JHU alone (p = 0.01 and p < 0.001, respectively; FWE for contrasts. Figure 4a, bottom row).

### 5) Secondary outcome 2: effect of psychosis type

We did not find a significant effect of psychosis type on group differences (Supplemental Results).

The regression coefficient between the PANSS positive scale and the mean cluster value of the MTLN was significantly larger in affective than in non-affective psychosis patients, both for all sites and JHU alone (p = 0.003 and p < 0.001, respectively; FWE for contrasts. Figure 4b, top row).

The regression coefficient between PANSS and the connectivity between the MTLN and BGN-MbThal was also significantly larger in affective than in non-affective psychosis patients for all sites and JHU alone (p = 0.001 and p < 0.001, respectively; FWE for contrasts. Figure 4b, bottom row).

### 6) Sensitivity analyses of the main and secondary outcomes

While a patient with the highest connectivity value in Figure 3 may appear to be an outlier, Cook’s distance (<0.5) indicated that this was not the case, and our permutation-based non-parametric inference was robust against outliers. However, we performed two sensitivity analyses: 1) excluding this participant and 2) using Spearman’s rank correlation (details in the Supplemental Results).

Excluding this patient did not affect the correlations with PANSS. The effect of medication on the within- and between-network correlation analyses showed trend-level significance, whereas the effects of psychosis type disappeared (all FWE for contrasts).

Spearman’s rank correlation revealed that the within- and between-network correlations with PANSS and the effect of medication on the between-network correlation remained significant, while other secondary analyses were not significant (all two-tailed).

### 7) Effects of comorbidity, smoking, and substance use

Comorbidity, smoking, and substance use did not affect any of the significant group difference results (Supplemental Results).

The effect of comorbidity on the regression coefficients between the PANSS positive scale and the connectivity of the MTLN showed a trend-level significance for within-network connectivity (Supplemental Figure S7) and fully corrected significance for between-network connectivity (Figure 4c) (p = 0.08 and 0.03, respectively; FWE for contrasts). Sub-analysis of the latter showed a trend-level positive correlation in patients without comorbidity and no correlation in patients with comorbidity (p = 0.06 and 0.279, respectively; FWE for contrasts). These findings indicated that the correlational findings were specific to psychosis rather than common among psychiatric disorders^64^.

Smoking showed trend-level effects on the correlations between PANSS and within- and between-connectivity of the MTLN (Supplemental Results and Supplemental Figure S8).

Substance use did not affect the correlation between connectivity and PANSS scores (Supplemental Results).

## Discussion

The main findings of this study were:

1: Reduced within-network connectivity and increased instability of the two salience-related systems in UHR and FEP, with the SN showing significance levels and effect sizes in the order UHR > FEP > ChrP.
2: Association between severer positive symptoms and stronger connectivity within and between the midbrain-striatum-hippocampus system in FEP.
3: Attenuation of finding 2 in medicated or non-affective psychosis patients. Collectively, we found fully corrected significance in UHR and FEP but not in ChrP, indicating that aberrant salience is more “salient” in early stages of psychosis^65^.

### Salience-related functional measures represent psychosis traits/staging

Regarding the midbrain-striatum-hippocampus system, we observed reduced connectivity in the thalamus of the BGN in patients with FEP. We recently reported heightened resting connectivity between the thalamus and cortical areas in patients with FEP^66^ in a sample that partially overlapped with the present study. Moreover, the thalamus has recently been reported to process salience^67^. Thus, the current results extend our knowledge of the underlying structure of aberrant salience in psychosis. In contrast, we did not observe altered within-network connectivity in the striatum of the BGN, whereas two previous ICA studies reported increased within-network connectivity^68,69^. Compared to these studies, we thoroughly considered confounding factors and strongly controlled for multiple comparisons.

For the ACC-insular SN system, we observed reduced within-network connectivity, with significance levels and effect sizes in the order of UHR > FEP > ChrP, in almost the same locations in the ACC (Figure 2b). This finding indicates that SN alterations are present at an early stage and may be weakened by chronicity. These findings are also largely consistent with those of previous ICA studies reporting reduced within-network SN connectivity in patients with schizophrenia ^70–74^.

This is the first ELA study to reveal frequent transitions between two brain states; that is, increased instability of salience-related networks in FEP. Previous studies have reported increased instability in the SN, FPN, and DMN in schizophrenia, consistent with our findings^75,76^. The finding of synchronized activation/deactivation patterns of all five NOIs in the two brain states is consistent with our view that the integration of different salience systems plays a key role in the pathophysiology of psychosis^77^.

Importantly, these findings were not affected by medication, psychosis type, comorbidities, smoking, or substance use. Therefore, the within-network connectivity and brain-state transition of the two salience-related systems are candidates for early-stage markers of psychosis; moreover, the ACC-insular SN system may also serve as a trait marker.

### Midbrain-striatum-hippocampus functional connectivity represents a psychosis state

We observed significant associations between more severe positive symptoms and stronger functional connectivity within the MTLN and between the MTLN and BGN-MbThal only at the FEP stage. These findings are consistent with the current view that pathophysiological changes are most drastic at this stage^37,38^, especially in the hippocampus^78^. To date, no study has investigated the MTLN in psychosis using rsfMRI and ICA; thus, our findings are not only new but also important, as the hippocampus is postulated to be the starting point of dopaminergic aberrant salience in psychosis^14^. The midbrain region of the BGN-MbThal was also associated with positive symptoms, although at a liberal threshold (Supplementary Figure S6a).

Importantly, these associations were attenuated in medicated patients, consistent with the requirements for state markers^34,35^. Moreover, this association was stronger in patients with affective psychosis than in those with non-affective psychosis. Since affective psychosis has better treatment responsiveness than non-affective psychosis^79^, these findings may represent responsiveness to antipsychotic medications. The finding that these associations were weakened by comorbidity and possibly smoking is not contradictory to this state-marker view.

We did not observe an association between positive symptoms and connectivity of the ACC-insular SN system at a fully corrected level, although we did observe associations at liberal thresholds (Supplementary Figure S6b). Taken together, these findings suggest that the functional connectivity of the midbrain-striatum-hippocampus system is a candidate state marker for aberrant salience, more so than the ACC-insular SN system.

### Robustness of the findings

A recent study reported that standard rsfMRI analysis with non-individualized cortical parcellation, without denoising, and without site/scanner-effect harmonization would require thousands of participants to attain statistical significance^80^. Our current findings, using ICA-based denoising, robust estimation of individualized large-scale networks by meta-ICA and thresholded dual regression, and harmonization by ComBat, were robust against full multiple comparison corrections. ComBat also efficiently harmonized the imbalance of medication status and psychosis type among our data, consistent with a previous report^59^.

The sensitivity analyses may indicate that the effect of medication is more robust than that of psychosis type or may be due to the smaller sample size in the latter (20 for unmedicated status while 13 for affective psychosis).

### Limitations

This study has several limitations. First, owing to the relative difficulties in recruiting UHR individuals and organizing a multi-site study, the UHR sample size was smaller than that of the FEP and ChrP. Second, susceptibility-induced distortion correction was not performed because several sites did not have the field maps required for this procedure. Third, while a longitudinal study design is more suitable, covering all stages is unrealistic, as it would require at least10 years of follow-up using the same scanner. Finally, developing clinically usable biomarkers from our findings will require their translation into more standardized frameworks, such as the estimation of individualized networks using the Bayesian approach with big-data population priors^81^, individualized cortical parcellation^82^, and/or combination with other modalities^66,83–85^.

### Conclusions

Functional alterations of the two salience-related systems, the midbrain-striatum-hippocampus and ACC-insular SN systems, were mainly associated with the early stages of psychosis. Alteration of the latter might be associated with psychosis traits, while the former was associated with the psychosis state, reflecting both positive symptoms and antipsychotic treatment. Further refinement of these findings will help to realize precision psychiatry. Our results provide a novel concept, stating that the two salience mechanisms work in both integrated and separate manners in psychosis.

## Data Availability

Data may be provided upon reasonable request after MTA.

## Acknowledgments

The authors thank all the participants in this study. The authors declare that they have no conflicts of interest regarding the submission of this manuscript, related to financial support or relationships.

This work was supported by:

The Japan Society for the Promotion of Science/Ministry of Education, Culture, Sports, Science and Technology (grant numbers KAKENHI JP26461767, JP17H04248, JP18H05130, JP20H05064, JP19H03583, JP23120009, JP16H06572, JP20K21567, JP15H04893, JP16H06397, JP16H06280, JP18K07550, and JP21K12153); the Japan Science and Technology Agency (grant numbers ImPACT JP15808865 and Moonshot R&D JPMJMS2021); the Japan Agency for Medical Research and Development Strategic Research Program for Brain Sciences (grant numbers JP17dm0107044 and JP20dm0107088), Brain/MINDS & beyond (grant numbers JP21dm0307008, JP20dm0307102, JP21dm0307001, JP21dm020769, JP21dm0307004, and JP21dm0307002), Brain Mapping by Integrated Neurotechnologies for Disease Studies (grant numbers JP20dm0207069, JP21dk0307103, and JP21uk1024002); and Intramural Research Grant (3-1) for Neurological and Psychiatric Disorders of the National Center of Neurology and Psychiatry; grants from the Japan Foundation for Aging and Health, a Novartis Pharma Research Grant, the SENSHIN Medical Research Foundation, the SUZUKEN Memorial Foundation, a Tanabe-Mitsubishi Pharma Research Grant, the Uehara Memorial Foundation, a Kyoto University Global Frontier Project for Young Professionals, the Takeda Science Foundation, the Kobayashi Magobei Memorial Foundation, the Smoking Research Foundation, and the Kato Memorial Trust for Nambyo Research; the National Institute of Mental Health (NIMH) (grant numbers MH-094268, MH-092443, and MH-105660), and the Silvio O. Conte Center funded by the NIMH; the National Institute of Health (grant numbers P41EB015909 and R01NS084957); and grants from Stanley, S-R/RUSK, and the National Alliance for Research on Schizophrenia and Depression.

Some of the participant recruitment was supported by Tanabe Mitsubishi Pharm. Co. Ltd; and a Wellcome Trust fellowship (grant number WT087779MA).

## References

1. Pearce JM, Hall G. A model for Pavlovian learning: Variations in the effectiveness of conditioned but not of unconditioned stimuli. Psychol Rev. 1980;87(6):532–552. doi:10.1037/0033-295X.87.6.532

2. Koch C, Ullman S. Shifts in selective visual attention: towards the underlying neural circuitry. Hum Neurobiol. 1985;4(4):219–227.

3. Matsumoto M, Hikosaka O. Two types of dopamine neuron distinctly convey positive and negative motivational signals. Nature. 2009;459(7248):837–841. doi:10.1038/nature08028

4. Matsumoto M, Takada M. Distinct Representations of Cognitive and Motivational Signals in Midbrain Dopamine Neurons. Neuron. 2013;79(5):1011–1024. doi:10.1016/j.neuron.2013.07.002

5. Menegas W, Akiti K, Amo R, Uchida N, Watabe-Uchida M. Dopamine neurons projecting to the posterior striatum reinforce avoidance of threatening stimuli. Nat Neurosci. 2018;21(10):1421–1430. doi:10.1038/s41593-018-0222-1

6. Zink CF, Pagnoni G, Martin-Skurski ME, Chappelow JC, Berns GS. Human Striatal Responses to Monetary Reward Depend On Saliency. Neuron. 2004;42(3):509–517. doi:10.1016/S0896-6273(04)00183-7

7. Zink CF, Pagnoni G, Chappelow J, Martin-Skurski M, Berns GS. Human striatal activation reflects degree of stimulus saliency. NeuroImage. 2006;29(3):977–983. doi:10.1016/j.neuroimage.2005.08.006

8. Jensen J, Smith AJ, Willeit M, et al. Separate brain regions code for salience vs. valence during reward prediction in humans. Hum Brain Mapp. 2007;28(4):294–302. doi:10.1002/hbm.20274

9. Cooper JC, Knutson B. Valence and salience contribute to nucleus accumbens activation. NeuroImage. 2008;39(1):538–547. doi:10.1016/j.neuroimage.2007.08.009

10. Kapur S. Psychosis as a State of Aberrant Salience: A Framework Linking Biology, Phenomenology, and Pharmacology in Schizophrenia. Am J Psychiatry. 2003;160(1):13–23. doi:10.1176/appi.ajp.160.1.13

11. Onitsuka T, Hirano Y, Nakazawa T, et al. Toward recovery in schizophrenia: Current concepts, findings, and future research directions. Psychiatry Clin Neurosci. 2022;76(7):282–291. doi:10.1111/pcn.13342

12. Fusar-Poli P, Meyer-Lindenberg A. Striatal Presynaptic Dopamine in Schizophrenia, Part II: Meta-Analysis of [18F/11C]-DOPA PET Studies. Schizophr Bull. 2013;39(1):33–42. doi:10.1093/schbul/sbr180

13. Crow TJ. Positive and Negative Schizophrenic Symptoms and the Role of Dopamine. Br J Psychiatry. 1980;137(4):383–386. doi:10.1192/S0007125000071919

14. Lodge DJ, Grace AA. Hippocampal dysregulation of dopamine system function and the pathophysiology of schizophrenia. Trends Pharmacol Sci. 2011;32(9):507–513. doi:10.1016/j.tips.2011.05.001

15. Winton-Brown T, Schmidt A, Roiser JP, et al. Altered activation and connectivity in a hippocampal–basal ganglia–midbrain circuit during salience processing in subjects at ultra high risk for psychosis. Transl Psychiatry. 2017;7(10):e1245. doi:10.1038/tp.2017.174

16. Modinos G, Allen P, Zugman A, et al. Neural Circuitry of Novelty Salience Processing in Psychosis Risk: Association With Clinical Outcome. Schizophr Bull. 2020;46(3):670–679. doi:10.1093/schbul/sbz089

17. Seeley WW, Menon V, Schatzberg AF, et al. Dissociable Intrinsic Connectivity Networks for Salience Processing and Executive Control. J Neurosci. 2007;27(9):2349–2356. doi:10.1523/JNEUROSCI.5587-06.2007

18. Dosenbach NUF, Fair DA, Miezin FM, et al. Distinct brain networks for adaptive and stable task control in humans. Proc Natl Acad Sci. 2007;104(26):11073–11078. doi:10.1073/pnas.0704320104

19. White TP, Joseph V, Francis ST, Liddle PF. Aberrant salience network (bilateral insula and anterior cingulate cortex) connectivity during information processing in schizophrenia. Schizophr Res. 2010;123(2-3):105–115. doi:10.1016/j.schres.2010.07.020

20. Palaniyappan L, Simmonite M, White TP, Liddle EB, Liddle PF. Neural Primacy of the Salience Processing System in Schizophrenia. Neuron. 2013;79(4):814–828. doi:10.1016/j.neuron.2013.06.027

21. Dong D, Wang Y, Chang X, Luo C, Yao D. Dysfunction of Large-Scale Brain Networks in Schizophrenia: A Meta-analysis of Resting-State Functional Connectivity. Schizophr Bull. 2018;44(1):168–181. doi:10.1093/schbul/sbx034

22. Palaniyappan L, Liddle PF. Does the salience network play a cardinal role in psychosis? An emerging hypothesis of insular dysfunction. J Psychiatry Neurosci. 2012;37(1):17–27. doi:10.1503/jpn.100176

23. Lavigne KM, Menon M, Woodward TS. Functional Brain Networks Underlying Evidence Integration and Delusions in Schizophrenia. Schizophr Bull. 2020;46(1):175–183. doi:10.1093/schbul/sbz032

24. Bressler SL, Menon V. Large-scale brain networks in cognition: emerging methods and principles. Trends Cogn Sci. 2010;14(6):277–290. doi:10.1016/j.tics.2010.04.004

25. Menon V, Uddin LQ. Saliency, switching, attention and control: a network model of insula function. Brain Struct Funct. 2010;214(5-6):655–667. doi:10.1007/s00429-010-0262-0

26. Hogeveen J, Krug MK, Elliott MV, Solomon M. Insula-Retrosplenial Cortex Overconnectivity Increases Internalizing via Reduced Insight in Autism. Biol Psychiatry. 2018;84(4):287–294. doi:10.1016/j.biopsych.2018.01.015

27. Bertocci MA, Afriyie-Agyemang Y, Rozovsky R, et al. Altered patterns of central executive, default mode and salience network activity and connectivity are associated with current and future depression risk in two independent young adult samples. Mol Psychiatry. December 2022:1–11. doi:10.1038/s41380-022-01899-8

28. Sha Z, Wager TD, Mechelli A, He Y. Common Dysfunction of Large-Scale Neurocognitive Networks Across Psychiatric Disorders. Biol Psychiatry. 2019;85(5):379–388. doi:10.1016/j.biopsych.2018.11.011

29. Supekar K, Cai W, Krishnadas R, Palaniyappan L, Menon V. Dysregulated Brain Dynamics in a Triple-Network Saliency Model of Schizophrenia and Its Relation to Psychosis. Biol Psychiatry. 2019;85(1):60–69. doi:10.1016/j.biopsych.2018.07.020

30. Robinson S, Basso G, Soldati N, et al. A resting state network in the motor control circuit of the basal ganglia. BMC Neurosci. 2009;10:137. doi:10.1186/1471-2202-10-137

31. Shirer WR, Ryali S, Rykhlevskaia E, Menon V, Greicius MD. Decoding Subject-Driven Cognitive States with Whole-Brain Connectivity Patterns. Cereb Cortex. 2012;22(1):158–165. doi:10.1093/cercor/bhr099

32. Meda SA, Gill A, Stevens MC, et al. Differences in Resting-State Functional Magnetic Resonance Imaging Functional Network Connectivity Between Schizophrenia and Psychotic Bipolar Probands and Their Unaffected First-Degree Relatives. Biol Psychiatry. 2012;71(10):881–889. doi:10.1016/j.biopsych.2012.01.025

33. Ritsner MS, Gottesman II. Where Do We Stand in the Quest for Neuropsychiatric Biomarkers and Endophenotypes and What Next? In: Ritsner MS, ed. *The Handbook of Neuropsychiatric Biomarkers*, Endophenotypes and Genes. Dordrecht: Springer Netherlands; 2009:3–21. doi:10.1007/978-1-4020-9464-4_1

34. Khoury R, Nasrallah HA. Inflammatory biomarkers in individuals at clinical high risk for psychosis (CHR-P): State or trait? Schizophr Res. 2018;199:31–38. doi:10.1016/j.schres.2018.04.017

35. Lema YY, Gamo NJ, Yang K, Ishizuka K. Trait and state biomarkers for psychiatric disorders: Importance of infrastructure to bridge the gap between basic and clinical research and industry. Psychiatry Clin Neurosci. 2018;72(7):482–489. doi:https://doi.org/10.1111/pcn.12669

36. National Institute of Mental Health Strategic Plan | 2008. :46.

37. Lieberman JA, First MB. Psychotic Disorders. N Engl J Med. 2018;379(3):270–280. doi:10.1056/NEJMra1801490

38. Howes OD, Hird EJ, Adams RA, Corlett PR, McGuire P. Aberrant Salience, Information Processing, and Dopaminergic Signaling in People at Clinical High Risk for Psychosis. Biol Psychiatry. 2020;88(4):304–314. doi:10.1016/j.biopsych.2020.03.012

39. Sakoğlu Ü, Pearlson GD, Kiehl KA, Wang YM, Michael AM, Calhoun VD. A method for evaluating dynamic functional network connectivity and task-modulation: application to schizophrenia. Magn Reson Mater Phys Biol Med. 2010;23(5):351–366. doi:10.1007/s10334-010-0197-8

40. Hutchison RM, Womelsdorf T, Allen EA, et al. Dynamic functional connectivity: Promise, issues, and interpretations. NeuroImage. 2013;80:360–378. doi:10.1016/j.neuroimage.2013.05.079

41. Aso T, Jiang G, Urayama S ichi, Fukuyama H. A Resilient, Non-neuronal Source of the Spatiotemporal Lag Structure Detected by BOLD Signal-Based Blood Flow Tracking. Front Neurosci. 2017;11. doi:10.3389/fnins.2017.00256

42. Roalf DR, Quarmley M, Elliott MA, et al. The impact of quality assurance assessment on diffusion tensor imaging outcomes in a large-scale population-based cohort. NeuroImage. 2016;125:903–919. doi:10.1016/j.neuroimage.2015.10.068

43. Smith SM, Fox PT, Miller KL, et al. Correspondence of the brain’s functional architecture during activation and rest. Proc Natl Acad Sci. 2009;106(31):13040–13045. doi:10.1073/pnas.0905267106

44. Biswal BB, Mennes M, Zuo XN, et al. Toward discovery science of human brain function. Proc Natl Acad Sci. 2010;107(10):4734–4739. doi:10.1073/pnas.0911855107

45. Poppe AB, Wisner K, Atluri G, Lim KO, Kumar V, MacDonald AW. Toward a neurometric foundation for probabilistic independent component analysis of fMRI data. Cogn Affect Behav Neurosci. 2013;13(3):641–659. doi:10.3758/s13415-013-0180-8

46. Bijsterbosch JD, Beckmann CF, Woolrich MW, Smith SM, Harrison SJ. The relationship between spatial configuration and functional connectivity of brain regions revisited. Ivry RB, Honey C, Margulies DS, Seidlitz J, eds. eLife. 2019;8:e44890. doi:10.7554/eLife.44890

47. Filippini N, MacIntosh BJ, Hough MG, et al. Distinct patterns of brain activity in young carriers of the APOE-ε4 allele. Proc Natl Acad Sci. 2009;106(17):7209–7214. doi:10.1073/pnas.0811879106

48. Watanabe T, Hirose S, Wada H, et al. A pairwise maximum entropy model accurately describes resting-state human brain networks. Nat Commun. 2013;4:1370. doi:10.1038/ncomms2388

49. Watanabe T, Hirose S, Wada H, et al. Energy landscapes of resting-state brain networks. Front Neuroinformatics. 2014;8. doi:10.3389/fninf.2014.00012

50. Watanabe T, Masuda N, Megumi F, Kanai R, Rees G. Energy landscape and dynamics of brain activity during human bistable perception. Nat Commun. 2014;5:4765. doi:10.1038/ncomms5765

51. Ezaki T, Watanabe T, Ohzeki M, Masuda N. Energy landscape analysis of neuroimaging data. Phil Trans R Soc A. 2017;375(2096):20160287. doi:10.1098/rsta.2016.0287

52. Watanabe T, Rees G. Brain network dynamics in high-functioning individuals with autism. Nat Commun. 2017;8:16048. doi:10.1038/ncomms16048

53. Ezaki T, Sakaki M, Watanabe T, Masuda N. Age-related changes in the ease of dynamical transitions in human brain activity. Hum Brain Mapp. 2018;39(6):2673–2688. doi:10.1002/hbm.24033

54. Onitsuka T, Hirano Y, Nemoto K, et al. Trends in big data analyses by multicenter collaborative translational research in psychiatry. Psychiatry Clin Neurosci. 2022;76(1):1–14. doi:10.1111/pcn.13311

55. Yamashita A, Yahata N, Itahashi T, et al. Harmonization of resting-state functional MRI data across multiple imaging sites via the separation of site differences into sampling bias and measurement bias. PLOS Biol. 2019;17(4):e3000042. doi:10.1371/journal.pbio.3000042

56. Yoshihara Y, Lisi G, Yahata N, et al. Overlapping but Asymmetrical Relationships Between Schizophrenia and Autism Revealed by Brain Connectivity. Schizophr Bull. 2020;46(5):1210–1218. doi:10.1093/schbul/sbaa021

57. Mori Y, Miyata J, Isobe M, et al. Effect of phase-encoding direction on group analysis of resting-state functional magnetic resonance imaging. Psychiatry Clin Neurosci. 2018;72(9):683–691. doi:10.1111/pcn.12677

58. Johnson WE, Li C, Rabinovic A. Adjusting batch effects in microarray expression data using empirical Bayes methods. Biostatistics. 2007;8(1):118–127. doi:10.1093/biostatistics/kxj037

59. Fortin JP, Parker D, Tunç B, et al. Harmonization of multi-site diffusion tensor imaging data. NeuroImage. 2017;161:149–170. doi:10.1016/j.neuroimage.2017.08.047

60. Yu M, Linn KA, Cook PA, et al. Statistical harmonization corrects site effects in functional connectivity measurements from multi-site fMRI data. Hum Brain Mapp. 2018;39(11):4213–4227. doi:https://doi.org/10.1002/hbm.24241

61. Winkler AM, Webster MA, Brooks JC, Tracey I, Smith SM, Nichols TE. Non-parametric combination and related permutation tests for neuroimaging: NPC and Related Permutation Tests for Neuroimaging. Hum Brain Mapp. 2016;37(4):1486–1511. doi:10.1002/hbm.23115

62. Suurmond R, van Rhee H, Hak T. Introduction, comparison, and validation of Meta-Essentials: A free and simple tool for meta-analysis. Res Synth Methods. 2017;8(4):537–553. doi:10.1002/jrsm.1260

63. Kay SR, Fiszbein A, Opler LA. The positive and negative syndrome scale (PANSS) for schizophrenia. Schizophr Bull. 1987;13(2):261–276.

64. Goodkind M, Eickhoff SB, Oathes DJ, et al. Identification of a Common Neurobiological Substrate for Mental Illness. JAMA Psychiatry. 2015;72(4):305–315. doi:10.1001/jamapsychiatry.2014.2206

65. Winton-Brown TT, Fusar-Poli P, Ungless MA, Howes OD. Dopaminergic basis of salience dysregulation in psychosis. Trends Neurosci. 2014;37(2):85–94. doi:10.1016/j.tins.2013.11.003

66. Faria AV, Zhao Y, Ye C, et al. Multimodal MRI assessment for first episode psychosis: A major change in the thalamus and an efficient stratification of a subgroup. Hum Brain Mapp. 2021;42(4):1034–1053. doi:https://doi.org/10.1002/hbm.25276

67. Zhu Y, Nachtrab G, Keyes PC, Allen WE, Luo L, Chen X. Dynamic salience processing in paraventricular thalamus gates associative learning. Science. 2018;362(6413):423–429. doi:10.1126/science.aat0481

68. Sorg C, Manoliu A, Neufang S, et al. Increased Intrinsic Brain Activity in the Striatum Reflects Symptom Dimensions in Schizophrenia. Schizophr Bull. 2013;39(2):387–395. doi:10.1093/schbul/sbr184

69. Duan M, Chen X, He H, et al. Altered basal ganglia network integration in schizophrenia. Front Hum Neurosci. 2015:561. doi:10.3389/fnhum.2015.00561

70. Manoliu A, Riedl V, Doll A, Bäuml JG, Bäuml J, Koch K. Insular dysfunction reflects altered between-network connectivity and severity of negative symptoms in schizophrenia during psychotic remission. Front Hum Neurosci. 2013;7:216. doi:10.3389/fnhum.2013.00216

71. Manoliu A, Riedl V, Zherdin A, et al. Aberrant Dependence of Default Mode/Central Executive Network Interactions on Anterior Insular Salience Network Activity in Schizophrenia. Schizophr Bull. 2014;40(2):428–437. doi:10.1093/schbul/sbt037

72. Wang Y, Tang W, Fan X, et al. Resting-state functional connectivity changes within the default mode network and the salience network after antipsychotic treatment in early-phase schizophrenia. Neuropsychiatr Dis Treat. 2017;Volume 13:397–406. doi:10.2147/NDT.S123598

73. Ohta M, Nakataki M, Takeda T, et al. Structural equation modeling approach between salience network dysfunction, depressed mood, and subjective quality of life in schizophrenia: an ICA resting-state fMRI study. Neuropsychiatric Disease and Treatment. doi:10.2147/NDT.S163132

74. Orliac F, Naveau M, Joliot M, et al. Links among resting-state default-mode network, salience network, and symptomatology in schizophrenia. Schizophr Res. 2013;148(1):74–80. doi:10.1016/j.schres.2013.05.007

75. Wang X, Zhang W, Sun Y, Hu M, Chen A. Aberrant intra-salience network dynamic functional connectivity impairs large-scale network interactions in schizophrenia. Neuropsychologia. 2016;93, Part A:262-270. doi:10.1016/j.neuropsychologia.2016.11.003

76. Supekar K, Cai W, Krishnadas R, Palaniyappan L, Menon V. Dysregulated Brain Dynamics in a Triple-Network Saliency Model of Schizophrenia and Its Relation to Psychosis. Biol Psychiatry. 2019;85(1):60–69. doi:10.1016/j.biopsych.2018.07.020

77. Miyata J. Toward integrated understanding of salience in psychosis. Neurobiol Dis. March 2019. doi:10.1016/j.nbd.2019.03.002

78. Goff DC, Zeng B, Ardekani BA, et al. Association of Hippocampal Atrophy With Duration of Untreated Psychosis and Molecular Biomarkers During Initial Antipsychotic Treatment of First-Episode Psychosis. JAMA Psychiatry. February 2018. doi:10.1001/jamapsychiatry.2017.4595

79. Demjaha A, Lappin JM, Stahl D, et al. Antipsychotic treatment resistance in first-episode psychosis: prevalence, subtypes and predictors. Psychol Med. 2017;47(11):1981–1989. doi:10.1017/S0033291717000435

80. Marek S, Tervo-Clemmens B, Calabro FJ, et al. Reproducible brain-wide association studies require thousands of individuals. Nature. March 2022:1–7. doi:10.1038/s41586-022-04492-9

81. Mejia AF, Nebel MB, Wang Y, Caffo BS, Guo Y. Template Independent Component Analysis: Targeted and Reliable Estimation of Subject-level Brain Networks using Big Data Population Priors. J Am Stat Assoc. 2020;115(531):1151–1177. doi:10.1080/01621459.2019.1679638

82. Glasser MF, Coalson TS, Robinson EC, et al. A multi-modal parcellation of human cerebral cortex. Nature. 2016;536(7615):171-178. doi:10.1038/nature18933

83. Wang AM, Pradhan S, Coughlin JM, et al. Assessing Brain Metabolism With 7-T Proton Magnetic Resonance Spectroscopy in Patients With First-Episode Psychosis. JAMA Psychiatry. January 2019. doi:10.1001/jamapsychiatry.2018.3637

84. Kamath V, Lasutschinkow P, Ishizuka K, Sawa A. Olfactory Functioning in First-Episode Psychosis. Schizophr Bull. 2018;44(3):672–680. doi:10.1093/schbul/sbx107

85. Faria AV, Crawford J, Ye C, et al. Relationship between neuropsychological behavior and brain white matter in first-episode psychosis. Schizophr Res. April 2019. doi:10.1016/j.schres.2019.04.010

