## Supplementary material for "Functional alterations of two salience-related systems jointly and independently contribute to psychosis"

**Supplemental Methods**

**Details of the participants**

We recruited 1) 29 individuals with ultra-high-risk for psychosis (UHR) and 25 age-, sex-, and ethnicity-matched healthy controls (HC); 2) 81 patients with first-episode psychosis (FEP) and 109 matched HC; and 3) 99 patients with chronic psychosis (ChrP) and 145 matched HC, comprising 488 subjects in total, from eight scanners at seven institutions. The details of each institution are described in the following sections 1) − 7) and Supplemental Tables S1 – S7. The Japanese institutions were members of the Cognitive Genetic Collaborative Research Organization (COCORO) consortium (http://www.sp-web.sakura.ne.jp/lab/cocoro.html).

UHR was defined using the Comprehensive Assessment of At-Risk Mental States (CAARMS)[1]. FEP and ChrP were defined as duration of illness of ≤ 36 months and > 36 months, respectively. Positive symptom severity was indexed according to the CAARMS "disorder of thought content" + "perceptual abnormalities" for UHR, and positive scale of the Positive and Negative Syndrome Scale (PANSS)[2] for FEP and ChrP. Chlorpromazine equivalent (CP eq) was calculated as described by Lehman et al[3] and Inada and Inagaki[4].

One individual with UHR from the Institute of Psychiatry, Psychology and Neuroscience (IoPPN) was medicated (50mg quetiapine). Johns Hopkins University (JHU) had both unmedicated (N=4) and medicated (N=47) patients with FEP, while other sites/scanners recruiting FEP and ChrP had either medication status alone.

Regarding psychosis type, JHU also had both affective (N=14, mood disorders with delusions/hallucinations) and non-affective (N=37, schizophrenia spectrum) patients with FEP, while the other sites had non-affective patients with FEP and ChrP only.

The HC individuals had no history of psychiatric illness or substance abuse. The exclusion criteria for all participants were a history of head trauma, neurological illness, or serious medical or surgical illness. The details of each site are as follows:

1) Institute of Psychiatry, Psychology and Neuroscience (IoPPN), UK

Participants with UHR and HC were recruited between December 2009 and September 2011. Individuals with UHR were referred to the IoPPN from the Outreach and Support in South London (OASIS) program. UHR was defined as the presentation of one or more of the following: 1) attenuated psychotic symptoms, 2) brief limited intermittent psychotic episodes, and/or 3) genetic risk and deterioration syndrome. The comorbidities in individuals with UHR included past or current depression (n = 13), personality disorders (n = 5), anxiety disorders (n = 5), and body dysmorphic disorder (n = 1). Among them, ten individuals with UHR had comorbidities other than schizophrenia spectrum or mood disorders. The CAARMS was used for diagnostic purposes in the intake assessment and when performing scans to assess symptom severity. The latter is presented in Table 1 of the main text. Individuals with HC had no first-degree relatives with psychotic disorders.

**There is no Supplemental Table for this site, because the demographic and clinical data are identical to those of the UHR sample of Table 1 in the main text.**

2) Johns Hopkins University (JHU), USA

Participants with FEP and HC were recruited between January 2014 and November 2015. The diagnosis was based on the Structured Clinical Interview for DSM-IV Axis I Disorders (SCID). FEP included schizophrenia, schizoaffective disorder, schizophreniform disorder, psychosis NOS, and depression or bipolar disorder with hallucinations or delusions. Six patients with FEP had comorbidities other than schizophrenia spectrum or mood disorders: childhood or current diagnosis of autism (n = 2), attention-deficit/hyperactivity disorder (n = 2), past history of anorexia nervosa (n = 1), and trichotillomania (n = 1). Symptomatology was assessed using the Scale for the Assessment of Positive Symptoms (SAPS)[5] and the Scale for the Assessment of Negative Symptoms (SANS)[6]. The SAPS global summary score was converted into the PANSS positive scale[7]. Forty-one patients were administered antipsychotics, four were unmedicated, and six were medicated but without information on the current dose. Participants in the HC group had no first-degree relatives with psychotic disorders. The demographic and clinical data are shown in Supplemental Table S1.

3) Kanazawa Medical University (KMU), Japan

Patients with FEP and ChrP as well as HCs were recruited between November 2011 and August 2018. All patients were diagnosed with schizophrenia using the SCID without comorbidity with other Axis I disorders. None of the patients were administered antipsychotics. The HCs had no first-degree relatives with psychotic disorders. The details are presented in Supplemental Table S2.

4) Kyoto University (Kyoto), Japan

Participants with ChrP and HC were recruited between October 2010 and June 2013. Ddiagnoses were made based on the SCID. ChrP included schizophrenia, schizoaffective disorder, and schizophreniform disorder. Patients were not comorbid with any other Axis I disorder. All patients were administered antipsychotics. The HCs had no first-degree relatives with psychotic disorders. The details are presented in Supplemental Table S3.

5) Two scanners from Osaka University (OSK2 and OSK3), Japan

Participants with FEP, ChrP, and HC were recruited between August 2011 and August 2013 (OSK2), and between February 2014 and January 2017 (OSK3). The diagnoses were confirmed using the SCID. All patients were diagnosed with schizophrenia, schizophreniform disorder, or brief psychotic disorder without comorbidity with other Axis I disorders. No patients were administered antipsychotics. The HCs had no first-degree relatives with psychotic disorders. The details are presented in Supplemental Tables S4 and S5.

6) The University of Tokyo (Tokyo), Japan
Patients with FEP and ChrP, as well as HC, were recruited between June 2014 and February 2019. All patients were diagnosed with schizophrenia without comorbidity with other Axis I disorders using the SCID. All the patients were administered antipsychotics. The details are presented in Supplemental Table S6.

7) University of Toyama (TYM), Japan
Patients with FEP and ChrP, as well as HC, were recruited between April 2015 and May 2017. All patients were diagnosed with schizophrenia without comorbidity with other Axis I disorders based on the SCID. None of the patients were administered antipsychotics. The HC subjects had no first-degree relatives with psychotic disorders. The details are presented in Supplemental Table S7.

**Magnetic resonance imaging (MRI) acquisition**

The sequences and parameters for each scanner are listed in Supplemental Table S8. All MRI images were visually checked, and participants with gross artifacts, anatomical anomalies, or excessive head motion (mean framewise displacement[8] of > 0.5 mm) were excluded from this study.

**Preprocessing of resting functional MRI (rsfMRI) data**

Head-motion correction was performed using SPM8 (Wellcome Department of Cognitive Neurology, London, United Kingdom) running on MATLAB (MathWorks), followed by spatial normalization to the MNI template brain using the T1 anatomical image.

Then, independent component analysis (ICA) denoising was performed to remove head motion, physiological noise such as pulsation and respiration, susceptibility-induced noise, and machine-derived noise[9]. Functional images were fed into MELODIC/FSL (http://www.fmrib.ox.ac.uk/fsl) with automatic independent component (IC) number estimation. Instead of using several participants as a training sample for machine-learning and classifying the ICs of other people into neuronal and noise components[10,11], we classified all participants' ICs based on three objective criteria: 1) high-frequency power ratio (>0.1 Hz up to 0.2 Hz or the Nyquist frequency for the repetition time [TR]) relative to <0.1 Hz, 2) non-gray matter involvement index, and 3) slice dependency index calculated as the ratio of within- and between-slice spatial high-frequency components. The thresholds of the three criteria for each site/scanner are presented in Supplemental Table S9. All 62 ICs of an example participant are shown in Supplemental Figure S1. ICs with a high-frequency power ratio (HF), high slice dependency index (SL), or high non-gray matter index (NB) were classified as noise, while components with neuronal origin were preserved. Each IC was interpreted according to a previously described machine-learning based protocol[10,11], showing consistency with our objective criteria. Supplemental video S1 also shows drastic noise reduction and temporal signal-to-noise ratio (tSNR) improvement by ICA denoising.

Denoised data were re-sampled into 3 mm isotropic voxels and smoothed by a Gaussian kernel of full width at half maximum of 5 mm. Because the minimum number of volumes was 150 (Supplemental Table S8), we used 145 volumes for each site/scanner and discarded the first five volumes for signal stabilization.

In this study, we did not perform temporal filtering (e.g., 0.01Hz – 0.1Hz); regarding low-pass (high-cut) filtering, neuronal signals are present beyond 0.1Hz up to at least 0.2Hz and there is no clear cutoff point[12]. For high-pass (low-cut) filtering, a current state-of-the-art preprocessing pipeline of the Human Connectome Project adopts a very unaggressive high-pass cutoff point of 0.0005Hz, casting a doubt on the necessity of high-pass filtering when ICA-denoising is performed[13]. In line with this, power spectra analysis of our data showed that ICA-denoising drastically removed low-frequency noises around 0.01Hz or slower (Supplemental Figure S2).

**Meta-ICA**

Because ICA algorithms are probabilistic, the networks identified by two ICA on the same data can differ slightly. To increase the robustness and reproducibility of group-level IC identification, we performed meta-ICA using the following steps.

1) First step: Five patients (PT) and five HC were randomly selected from each site/scanner. The preprocessed rsfMRI data of 80 participants were temporarily concatenated, and group ICA (gICA) was performed using the MELODIC[14] toolbox of FSL (version 6.0.3) (http://www.fmrib.ox.ac.uk/fsl). This step was iterated 25 times to attain robustness of ICs[15].

2) Second step: The results (i.e., ICs) of 25 gICA were concatenated, and a “grand” gICA was performed. The results were thresholded at $p > 0.5$, using alternative hypothesis testing in a Gaussian/Gamma mixture model.

We repeated these steps incrementing the total IC number from 20 by 5. At an IC total of 80, the networks of interest (NOIs: the basal ganglia network [BGN], salience network [SN] and medial temporal lobe network [MTLN]) were identified by visual inspection and confirmed by matching to templates, as described in the next section.

**Network identification**

After visual inspection, NOIs were confirmed by matching them to templates for the BGN, SN, and MTLN. The templates were created as follows:

1) BGN template: The bilateral thalamus, caudate, putamen, and accumbens were extracted from the Harvard-Oxford subcortical structural atlas in FSL (HarvardOxford-sub-maxprob-thr25-2mm), binarized, combined, and resliced into 3 mm isotropic voxels (Supplemental Figure S3a).

2) SN template: The anterior cingulate cortex (ACC) and bilateral insula were extracted from the atlas above, binarized, combined, and resliced into 3 mm voxels (Supplemental Figure S3b).

3) MTLN template: The bilateral amygdala, hippocampus, and anterior/posterior parahippocampal gyrus were extracted from the same atlas, binarized, combined, and then resliced (Supplemental Figure S3c).

Then, all 80 ICs were cross correlated with these three templates using the fslcc command in FSL. Supplemental Table S10 shows that the five visually identified NOIs had the highest correlation coefficients with each template among the 80 ICs.

**Thresholded dual regression**

Thresholded dual regression[16] in FSL comprised four steps, in which the first and second steps were to obtain individualized spatial maps for each IC and the third and fourth steps were to provide individualized time-courses.

First, according to the conventional dual regression procedure[17], we regressed each subject's preprocessed data against the meta-ICA maps created above to obtain subject-specific time-courses for each IC. Second, we regressed the preprocessed data against these subject-specific time-courses to obtain subject-specific spatial maps for each IC. Third, Gaussian-Gamma mixture model thresholding was applied to each subject's spatial maps[14]. Fourth, we regressed the preprocessed data against these thresholded maps to obtain subject-specific time-courses.

The time courses provided in the fourth step represented the regions inside the networks, whereas those of the first step represented the whole brain, including the regions outside the networks[16].

Finally, we calculated partial correlation coefficients between the time courses of NOI-pairs using FSLNets version 0.6.3, while controlling for the other three NOIs, and then Fisher transformed them into Z-scores.

**Dynamic analysis by energy landscape analysis (ELA)**

Using the ELA toolbox (https://github.com/tkEzaki/energy-landscape-analysis), each time point of the time courses of the five NOIs was binarized into 1 ("activated") and -1 ("deactivated") depending on whether the value of a time point was above or below the mean of each time course. Since we had five NOIs, the possible activity patterns at each time point were $2^5 = 32$ patterns. We concatenated the binarized time-series data of all the participants and calculated the frequency (probability) of each activity pattern. We then fitted the pairwise maximum entropy model to the data to calculate the "energy" of each activity pattern and classified the 32 activity patterns into two brain states, A and B. State A had a local minima (frequent, low-energy pattern) in which all five NOIs were synchronized in deactivation, whereas state B had a local minima in which all NOIs were activated in synchronization (Figure 1b and 1c).

For each subject, the frequency of state B and transition rate between states A and B were calculated. The frequency of state A was (1 minus [frequency of state B]).

**Site/scanner-effect harmonization by ComBat**

For within-network analysis, we modeled the additive (γ) and multiplicative (δ) site/scanner effects at each voxel using the following formula:

$$y_{ijv} = \alpha_v + \mathbf{X}_{ij}\boldsymbol{\beta}_v + \gamma_{iv} + \delta_{iv}\varepsilon_{ijv}$$

Where $y_{ijv}$ represents the value of voxel v, subject j, and site/scanner i=1,..., 8. α is the mean for voxel v, X is a design matrix including diagnosis, age, sex, and tSNR, β is the voxel-specific vector of regression coefficients corresponding to X, and ε is the error term. Medication, psychosis type, and comorbidities were not included in X because HC group lacked these variables. Similarly, smoking and substance use were not included in X, because they were not available at several sites.

All voxels were assumed to have the same distribution (γ – Gaussian and δ – inverse gamma distributions). The hyperparameters of these distributions were estimated empirically from the data. γ and δ were estimated using conditional posterior means and were finally removed from the voxel values using the following formula:

$$y_{ijv}^{ComBat} = \frac{y_{ijv} - \widehat{\alpha}_v - \mathbf{X}_{ij}\widehat{\boldsymbol{\beta}}_v - \gamma_{iv}^*}{\delta_{iv}^*} + \widehat{\alpha}_v + \mathbf{X}_{ij}\widehat{\boldsymbol{\beta}}_v$$

The same procedures were performed for the Z-transformed correlation coefficients of the between-network analysis and the frequency and transition rate of the ELA. Supplemental Figure S4 illustrates efficient removal of the site/scanner effect.

**Details of the statistical analysis**

Demographic and clinical data were analyzed using SPSS 27 (IBM). For imaging analyses, general linear model (GLM) was used for group comparisons and correlational analyses. Permutation-based non-parametric inference using PALM in FSL was performed[18], to produce the permutation distribution of the extremum statistic across all tests and compute family-wise error (FWE)-corrected p-values. Multiple comparisons were corrected for:

A) Voxels (within-network analysis only): threshold-free cluster enhancement[19] within each NOI.

B) Number of contrasts: two contrasts (HC > PT and PT > HC for group comparisons and positive or negative correlations for correlational analyses).

C) Number of networks/network pairs/ELA indices: five NOIs for within-network analysis, 10 NOI-pairs for between-network analysis, and two indices (frequency and transition rate) for ELA.

Control across the psychosis stages was not performed, since this study could be considered a series of independent studies at each stage.

**Procedures to investigate medication and psychosis-type effects**

1) For within-network connectivity analyses, the mean value of the significant clusters was extracted and used for the investigation. We used significant network measures for between-network and dynamic analyses.

2) For UHR, we performed group comparisons or correlational analyses after excluding one patient who was administered 50 mg quetiapine. For group comparisons of FEP and ChrP, we compared the above network values between unmedicated and medicated patients. For correlational analyses of FEP and ChrP, we compared the regression coefficients of unmedicated and medicated patients between positive symptom severity and network measures in the form of interaction analysis using GLM.

3) We examined the effect of psychosis type (affective or non-affective) for FEP similarly. Since all patients with ChrP had non-affective psychosis, we did not investigate this stage.

4) Age, sex, and tSNR were used as covariates. The statistical threshold was set at $p < 0.05$, FWE corrected for contrasts. If more than two networks/network pairs/ELA indices were found to be significant at each stage, we also corrected them.

**Effects of comorbidity, smoking, and substance use**

Ten individuals with UHR from IoPPN and six patients with FEP from JHU had comorbid psychiatric disorders in addition to schizophrenia spectrum or mood disorders, as described above. Thus, for these sites, we divided the patients into those with and without comorbidities and investigated the effect of comorbidity in the same way as medication and psychosis type.

Information on smoking was available from the IoPPN, JHU, Kyoto, OSK, and Tokyo. The effects of smoking were investigated in a similar manner.

Four individuals with UHR from IoPPN and 11 patients with FEP from JHU had experience with substance use, and its effects were investigated similarly.

### Supplemental Results

#### Demographic data

As shown in Table 1, the age, sex, ethnicity, and mean tSNR did not differ significantly between the PT and HC groups at any stage. The number of years of completed education was lower in the PT group than that in HC group in all stages. Smoking and substance use were more prevalent in patients with FEP than in HC.

#### No effects of medication on the group difference results

After excluding an individual with UHR administered a low dose of quetiapine (50 mg), the group comparison of the cluster value of the SN-ACC remained significant ($p < 0.001$, FWE corrected for contrasts).

For FEP, comparison between unmedicated and medicated patients revealed no significant difference in the cluster value of the right thalamus of the BGN-MbThal, both for all sites and JHU only ($p = 0.43$ and 0.41, respectively, uncorrected).

The transition rate between the two brain states did not differ between unmedicated and medicated patients ($p = 0.064$ and 0.28 for all sites and JHU only, respectively, uncorrected).

#### No effects of psychosis type on the group difference results

Comparison between affective and non-affective psychosis did not reveal significant differences in the cluster value of the right thalamus of the BGN-MbThal, both for all sites and JHU only ($p = 0.20$ and $p = 0.40$, respectively, uncorrected).

The transition rate between the two brain states did not differ between the psychosis types ($p = 0.066$ and $p = 0.15$ for all sites and JHU only, respectively, uncorrected).

#### Sensitivity analyses for the main and secondary outcomes

We performed sensitivity analyses by 1) excluding a patient with FEP with the highest connectivity value and 2) using Spearman's rank correlation coefficient.

1) Excluding one patient

In the main outcome, the correlation between the PANSS positive scale and within- and between-network connectivity of the MTLN remained significant ($p < 0.001$ and $p = 0.02$, respectively; FWE corrected for contrasts).

Regarding the secondary outcomes of the effect of medication, the regression coefficient between PANSS and the within- and between-network connectivity of the MTLN were larger in unmedicated than in medicated patients at trend-level significance ($p = 0.053$ and 0.06 for within- and between-network, respectively, FWE for contrasts). For psychosis type, the regression coefficient of the within- and between-network connectivity of the MTLN did not differ between affective and non-affective psychosis ($p = 0.23$ and 0.29 for within- and between-network, respectively, uncorrected).

2) Spearman's rank correlation analyses

We calculated rank correlation coefficients between the PANSS positive scale and the connectivity values (age, sex, tSNR were adjusted), and compared the Z-transformed correlation coefficients between medication status/psychosis type using SPSS. Statistical significance was set at $p < 0.05$ (two-tailed test).

The main outcome of the correlation between PANSS and within-network connectivity of the MTLN remained significant ($p < 0.001$, two-tailed).

In the secondary outcomes, the correlation coefficient between PANSS and the within-network connectivity of the MTLN did not differ between medication statuses ($p = 0.824$, two-tailed), while that of between-network connectivity of the MTLN was significantly larger in unmedicated than in medicated patients ($p = 0.023$, two-tailed). The correlation coefficients of the within- and between-network connectivity of the MTLN did not differ between psychosis types ($p = 0.7$ and 0.48, for within- and between-network respectively, two-tailed).

**Effect of comorbidity on the group difference results**

For UHR, the cluster value of the ACC of the SN-ACC did not differ between individuals with (n=10) and without (n=19) comorbidities ($p = 0.35$, uncorrected).

For FEP, the cluster value of the right thalamus of BGN-MbThal did not differ between patients with (n=6) and without (n=45) comorbidities ($p = 0.33$, uncorrected).

The transition rate of ELA in FEP did not differ between patients with and without comorbidities ($p = 0.2$, uncorrected).

**Effect of smoking on the group difference results**

For UHR, the cluster value of the ACC of the SN-ACC did not differ between non-smokers (n = 10) and smokers (n = 16) ($p = 0.27$, uncorrected).

For FEP, the cluster value of the right thalamus of the BGN-MbThal did not differ between non-smokers (n = 57) and smokers (n = 16) (p = 038, uncorrected).

The transition rate of ELA in FEP did not differ between non-smokers and smokers ($p = 0.41$, uncorrected).

**Effect of substance use on the group difference results**

For UHR, the cluster value of the ACC of the SN-ACC did not differ between non-users (25) and users (4) (p = 0.18, uncorrected).

For FEP, the cluster value of the right thalamus of the BGN-MbThal did not differ between non-users (n = 37) and users (n = 11) (p = 0.26, uncorrected).

The transition rate between the two brain states did not differ between non-users and users (p = 0.17, uncorrected).

**Effect of smoking on the correlational analysis**

The regression coefficients between the PANSS positive scale and the within- and between-network connectivity of the MTLN were larger in non-smokers (n=54) than in smokers (n=15) at the trend level (p = 0.07 and 0.052, for within- and between-network, respectively, FWE corrected for contrasts) (Supplemental Figure S8).

**Effect of substance use on the correlational analysis**

The regression coefficients between the PANSS positive scale and the within- and between-network connectivity of the MTLN did not differ between non-users (n=34) and users (n=10) (p = 0.096 and 0.064, for within- and between-network, respectively, uncorrected).

Supplemental references


1. Yung AR, Yuen HP, McGorry PD, et al. Mapping the onset of psychosis: the Comprehensive Assessment of At-Risk Mental States. *Aust N Z J Psychiatry*. 2005;39(11-12):964-971. doi:10.1080/j.1440-1614.2005.01714.x

2. Kay SR, Fiszbein A, Opler LA. The positive and negative syndrome scale (PANSS) for schizophrenia. *Schizophr Bull*. 1987;13(2):261-276.

3. Lehman AF, Lieberman JA, Dixon LB, et al. Practice guideline for the treatment of patients with schizophrenia, second edition. *Am J Psychiatry*. 2004;161(2 Suppl):1-56.

4. Inada T, Inagaki A. Psychotropic dose equivalence in Japan. *Psychiatry Clin Neurosci*. 2015;69(8):440-447. doi:10.1111/pcn.12275

5. Andreasen NC. *Scale for the Assessment of Positive Symptons: SAPS*. University of Iowa; 1984.

6. Andreasen NC. *Scale for the Assessment of Negative Symptoms*. University of Iowa; 1981.

7. van Erp TGM, Preda A, Nguyen D, et al. Converting positive and negative symptom scores between PANSS and SAPS/SANS. *Schizophrenia Research*. 2014;152(1):289-294. doi:10.1016/j.schres.2013.11.013

8. Jenkinson M, Bannister P, Brady M, Smith S. Improved Optimization for the Robust and Accurate Linear Registration and Motion Correction of Brain Images. *NeuroImage*. 2002;17(2):825-841. doi:10.1006/nimg.2002.1132

9. Aso T, Jiang G, Urayama S ichi, Fukuyama H. A Resilient, Non-neuronal Source of the Spatiotemporal Lag Structure Detected by BOLD Signal-Based Blood Flow Tracking. *Front Neurosci*. 2017;11. doi:10.3389/fnins.2017.00256

10. Griffanti L, Salimi-Khorshidi G, Beckmann CF, et al. ICA-based artefact removal and accelerated fMRI acquisition for improved resting state network imaging. *NeuroImage*. 2014;95(Supplement C):232-247. doi:10.1016/j.neuroimage.2014.03.034

11. Salimi-Khorshidi G, Douaud G, Beckmann CF, Glasser MF, Griffanti L, Smith SM. Automatic denoising of functional MRI data: Combining independent component analysis and

hierarchical fusion of classifiers. *NeuroImage*. 2014;90(Supplement C):449-468. doi:10.1016/j.neuroimage.2013.11.046

12. Niazy RK, Xie J, Miller K, Beckmann CF, Smith SM. Chapter 17 - Spectral characteristics of resting state networks. In: Van Someren EJW, Van Der Werf YD, Roelfsema PR, Mansvelder HD, Lopes Da Silva FH, eds. *Progress in Brain Research*. Vol 193. Slow Brain Oscillations of Sleep, Resting State and Vigilance. Elsevier; 2011:259-276. doi:10.1016/B978-0-444-53839-0.00017-X

13. Smith SM, Beckmann CF, Andersson J, et al. Resting-state fMRI in the Human Connectome Project. *NeuroImage*. 2013;80:144-168. doi:10.1016/j.neuroimage.2013.05.039

14. Beckmann CF, Smith SM. Probabilistic independent component analysis for functional magnetic resonance imaging. *IEEE Transactions on Medical Imaging*. 2004;23(2):137-152. doi:10.1109/TMI.2003.822821

15. Poppe AB, Wisner K, Atluri G, Lim KO, Kumar V, MacDonald AW. Toward a neurometric foundation for probabilistic independent component analysis of fMRI data. *Cogn Affect Behav Neurosci*. 2013;13(3):641-659. doi:10.3758/s13415-013-0180-8

16. Bijsterbosch JD, Beckmann CF, Woolrich MW, Smith SM, Harrison SJ. The relationship between spatial configuration and functional connectivity of brain regions revisited. Ivry RB, Honey C, Margulies DS, Seidlitz J, eds. *eLife*. 2019;8:e44890. doi:10.7554/eLife.44890

17. Filippini N, MacIntosh BJ, Hough MG, et al. Distinct patterns of brain activity in young carriers of the APOE-ε4 allele. *Proceedings of the National Academy of Sciences*. 2009;106(17):7209-7214. doi:10.1073/pnas.0811879106

18. Winkler AM, Webster MA, Brooks JC, Tracey I, Smith SM, Nichols TE. Non-parametric combination and related permutation tests for neuroimaging: NPC and Related Permutation Tests for Neuroimaging. *Human Brain Mapping*. 2016;37(4):1486-1511. doi:10.1002/hbm.23115

19. Smith SM, Nichols TE. Threshold-free cluster enhancement: Addressing problems of smoothing, threshold dependence and localisation in cluster inference. *NeuroImage*. 2009;44(1):83-98. doi:10.1016/j.neuroimage.2008.03.061

**Supplemental Table S1. Demographic and clinical data of individuals recruited at Johns Hopkins University (JHU)**

| | HC (73) | FEP (51) | Statistics | DoF | P |
|---|---|---|---|---|---|
| Age, mean years (SD) | 24.1 (3.7) | 22.6 (4.7) | t = 1.85 | 90.5 | 0.07 |
| Age range, years | 18-34 | 15-34 | | | |
| Sex (female/male) | 36/37 | 17/34 | Chi-square = 3.1 | 1 | 0.1* |
| Ethnicity | | | Chi-square = 2.7 | 5 | 0.75 |
| African | 43 | 26 | | | |
| Caucasian | 26 | 19 | | | |
| Asian | 2 | 3 | | | |
| Hispanic | 0 | 1 | | | |
| Mixed | 1 | 1 | | | |
| Unknown | 1 | 1 | | | |
| Education (year) | 14.7 (2.1) | 13.2 (3.1) | t = 3.0 | 82.1 | 0.004 |
| Smoking (yes/no) | 4/69 | 16/35 | Chi-square = 14.9 | 1 | < 0.001* |
| Cannabis use (yes/no) | 2/71 | 11/37 | Chi-square = 12.3 | 1 | 0.001* |
| Duration of illness (years) | | 1.3 (0.9) | | | |
| SAPS total | | 15.6 (20.0) | | | |
| SANS total | | 30.7 (21.9) | | | |
| PANSS positive ** | | 13.0 (4.2) | | | |
| CP eq. | | 470.1 (321.6) | | | |
| tSNR | 5065.2 (1563.0) | 5065.0 (1963.3) | t = 0.001 | 122 | 1.00 |

* Fisher's exact test.

** Converted from SAPS.

**Abbreviations**

CP eq, chlorpromazine equivalent; DoF, degree of freedom; FEP, first-episode psychosis; HC, healthy controls; PANSS, Positive and Negative Syndrome Scale; SANS, Scale for the Assessment of Negative Symptoms; SAPS, Scale for the Assessment of Positive Symptoms; tSNR, temporal signal/noise ratio.

**Supplemental Table S2. Demographic and clinical data of individuals recruited at Kanazawa Medical University (KMU)**

| | HC (15) | FEP (4) & ChrP (4) | Statistics | DoF | P |
|---|---|---|---|---|---|
| Age, mean years (SD) | 29.7 (8.8) | 32.0 (12.2) | t = -0.53 | 21 | 0.6 |
| Age range, years | 19-49 | 20-51 | | | |
| Sex (female/male) | 10/5 | 2/6 | Chi-square = 3.6 | 1 | 0.09* |
| Ethnicity | All Japanese | All Japanese | | | |
| Education (year) | 16.5 (1.6) | 13.1 (1.8) | t = 4.6 | 20 | < 0.001 |
| Smoking | Not available | Not available | | | |
| Substance use | All negative | All negative | | | |
| Duration of illness (years) | | 3.1 (3.7) | | | |
| PANSS positive | | 17.4 (4.9) | | | |
| PANSS negative | | 18.8 (5.7) | | | |
| PANSS general | | 30.1 (6.7) | | | |
| CP eq | | 0 | | | |
| tSNR | 1866.9 (373.6) | 2004.3 (493.5) | t = -0.75 | 21 | 0.46 |

* Fisher's exact test.

**Abbreviations**

ChrP, chronic schizophrenia; CP eq, chlorpromazine equivalent; DoF, degree of freedom; FEP, first-episode psychosis; HC, healthy controls; PANSS, Positive and Negative Syndrome Scale; tSNR, temporal signal/noise ratio.

**Supplemental Table S3. Demographic and clinical data of individuals recruited at Kyoto University (Kyoto)**

| | HC (41) | ChrP (46) | Statistics | DoF | P |
|---|---|---|---|---|---|
| Age, mean years (SD) | 36.3 (6.7) | 39.0 (9.2) | t = -1.57 | 82.1 | 0.12 |
| Age range, years | 30-57 | 24-57 | | | |
| Sex (female/male) | 11/30 | 21/25 | Chi-square = 3.3 | 1 | 0.08* |
| Ethnicity | All Japanese | All Japanese | | | |
| Education (years) | 16.7 (2.2) | 14.1 (2.2) | t = 5.26 | 81 | < 0.001 |
| Smoking (yes/no) | 5/31 | 9/34 | Chi-square = 0.67 | 1 | 0.56* |
| Substance use | All negative | All negative | | | |
| Duration of illness (years) | | 14.3 (7.8) | | | |
| PANSS positive | | 14.6 (5.1) | | | |
| PANSS negative | | 15.7 (5.6) | | | |
| PANSS general | | 29.6 (9.8) | | | |
| CP eq | | 591.8 (480.1) | | | |
| tSNR | 1423.0 (334.5) | 1681.7 (551.9) | t = -2.68 | 75.3 | 0.009 |

* Fisher's exact test

**Abbreviations**

ChrP, chronic schizophrenia; CP eq, chlorpromazine equivalent; DoF, degree of freedom; HC, healthy controls; PANSS, Positive and Negative Syndrome Scale; tSNR, temporal signal/noise ratio.

**Supplemental Table S4. Demographic and clinical data of individuals recruited at Osaka University MRI2 (OSK2)**

| | HC (17) | FEP (2) & ChrP (7) | Statistics | DoF | P |
|---|---|---|---|---|---|
| Age, mean years (SD) | 32.4 (15.4) | 33.6 (16.2) | t = -.186 | 24 | 0.85 |
| Age range, years | 20-64 | 20-64 | | | |
| Sex (female/male) | 12/5 | 6/3 | Chi-square = 0.042 | 1 | 1.00* |
| Ethnicity | All Japanese | All Japanese | | | |
| Education (years) | 14.7 (2.4) | 13.2 (1.9) | t = 1.54 | 24 | 0.14 |
| Smoking (yes/no) | (0/17) | (2/7) | | | |
| Substance use | All negative | All negative | | | |
| Duration of illness (years) | | 8.2 (7.3) | | | |
| PANSS positive | | 18.7 (4.9) | | | |
| PANSS negative | | 20.3 (5.9) | | | |
| PANSS general | | 45.4 (9.8) | | | |
| CP eq | | 0 | | | |
| tSNR | 7950.8 (1176.9) | 8559.7 (872.1) | t = -1.36 | 24 | 0.19 |

* Fisher's exact test

**Abbreviations**

ChrP, chronic schizophrenia; CP eq, chlorpromazine equivalent; DoF, degree of freedom; FEP, first-episode psychosis; HC, healthy controls; PANSS, Positive and Negative Syndrome Scale; tSNR, temporal signal/noise ratio.

**Supplemental Table S5. Demographic and clinical data of individuals recruited at Osaka University MRI3 (OSK3)**

| | HC (35) | FEP (7) & ChrP (14) | Statistics | DoF | P |
|---|---|---|---|---|---|
| Age, mean years (SD) | 30.5 (11.3) | 27.7 (8.0) | t = 1.01 | 54 | 0.32 |
| Age range, years | 18-66 | 16-43 | | | |
| Sex (female/male) | 19/16 | 11/10 | Chi-square = 0.19 | 1 | 1.00* |
| Ethnicity | All Japanese | All Japanese | | | |
| Education (years) | 14.9 (2.4) | 13.4 (2.7) | t = 2.15 | 54 | 0.04 |
| Smoking (yes/no) | 5/30 | 0/21 | | | |
| Substance use | All negative | All negative | | | |
| Duration of illness (years) | | 5.8 (5.6) | | | |
| PANSS positive | | 20.1 (4.2) | | | |
| PANSS negative | | 21.9 (4.3) | | | |
| PANSS general | | 47.2 (10.8) | | | |
| CP eq | | 0 | | | |
| tSNR | 17285.8 (2516.7) | 20959.8 (4610.9) | t = -3.36 | 27.3 | 0.002 |

* Fisher's exact test

**Abbreviations**

ChrP, chronic schizophrenia; CP eq, chlorpromazine equivalent; DoF, degree of freedom; FEP, first-episode psychosis; HC, healthy controls; PANSS, Positive and Negative Syndrome Scale; tSNR, temporal signal/noise ratio.

**Supplemental Table S6. Demographic and clinical data of individuals recruited at the University of Tokyo (Tokyo)**

| | HC (63) | FEP (14) & ChrP (26) | Statistics | DoF | P |
|---|---|---|---|---|---|
| Age, mean years (SD) | 38.7 (8.2) | 28.9 (10.0) | t = 5.43 | 101 | < 0.001 |
| Age range, years | 25-59 | 16-52 | | | |
| Sex (female/male) | 37/26 | 17/23 | Chi-square = 2.58 | 1 | 0.16* |
| Ethnicity | All Japanese | All Japanese | | | |
| Education (years) | 15.8 (2.3) | 13.4 (2.4) | t = 5.14 | 101 | < 0.001 |
| Smoking (yes/no) | 6/57 | 4/31 | Chi-square = 0.09 | 1 | 0.74* |
| Substance use | All negative | All negative | | | |
| Duration of illness (years) | | 8.6 (8.6) | | | |
| PANSS positive | | 16.2 (4.9) | | | |
| PANSS negative | | 19.0 (6.7) | | | |
| PANSS general | | 33.6 (7.7) | | | |
| CP eq | | 546.5 (431.3) | | | |
| tSNR | 19682.8 (4260.1) | 18862.8 (3209.5) | t = 1.04 | 101 | 0.3 |

* Fisher's exact test

**Abbreviations**

ChrP, chronic schizophrenia; CP eq, chlorpromazine equivalent; DoF, degree of freedom; FEP, first-episode psychosis; HC, healthy controls; PANSS, Positive and Negative Syndrome Scale; tSNR, temporal signal/noise ratio.

**Supplemental Table S7. Demographic and clinical data of individuals recruited at the University of Toyama (TYM)**

| | HC (10) | FEP (3) & ChrP (2) | Statistics | DoF | P |
|---|---|---|---|---|---|
| Age, mean years (SD) | 25.30 (5.67) | 28.20 (7.91) | t = -.82 | 13 | 0.43 |
| Age range, years | 21-37 | 21-39 | | | |
| Sex (female/male) | 6/4 | 2/3 | Chi-square = 0.54 | 1 | 0.61* |
| Ethnicity | All Japanese | All Japanese | | | |
| Education (years) | 16.5 (1.6) | 14.4 (1.7) | t = 2.38 | 13 | 0.03 |
| Smoking | Not available | Not available | | | |
| Substance use | All negative | All negative | | | |
| Duration of illness (years) | | 3.2 (3.8) | | | |
| PANSS positive | | 15.8 (3.6) | | | |
| PANSS negative | | 17.0 (7.6) | | | |
| PANSS general | | 35.8 (8.4) | | | |
| CP eq | | 0 | | | |
| tSNR | 2610.6 (340.8) | 2885.9 (512.3) | t = -1.25 | 13 | 0.23 |

* Fisher's exact test

**Abbreviations**

ChrP, chronic schizophrenia; CP eq, chlorpromazine equivalent; DoF, degree of freedom; FEP, first-episode psychosis; HC, healthy controls; PANSS, Positive and Negative Syndrome Scale; tSNR, temporal signal/noise ratio.

**Supplemental Table S8. Scanning parameters at each scanner/institution.**

| | IoPPN | JHU | KMU | Kyoto | OSK2 | OSK3 | Tokyo | TYM |
|---|---|---|---|---|---|---|---|---|
| Vendor | Siemens | Philips | Siemens | Siemens | GE | GE | GE | Siemens |
| Type | TimTRIO | Achieva | TimTRIO | TRIO | Signa HDxt | DISCOVERY MR750 | DISCOVERY MR750w | Verio |
| Head coil (channel) | 32 | 32 | 32 | 8 | 8 | 8 | 24 | 12 |
| Magnetic field | 3T (Tesla) | 3T | 3T | 3T | 3T | 3T | 3T | 3T |
| T1-weighted image | | | | | | | | |
| Sequence | MP2RAGE | MPRAGE | MPRAGE | MPRAGE | FSPGR | FSPGR | FSPGR | MPRAGE |
| Repetition time (ms) | 5000 | 6.8 | 1420 | 2000 | 7.2 | 8.16 | 7.65 | 2300 |
| Echo time (ms) | 2.96 | 3.1 | 2.08 | 4.38 | 2.9 | 3.17 | 3.1 | 2.9 |
| Inversion time (ms) | 700 & 2500 | 845 | 800 | 990 | 400 | 400 | 400 | 900 |
| Flip angle | 4 | 8 | 9 | 8 | 11 | 11 | 11 | 9 |
| Resolution (x/y/z in mm) | 1/1/1 | 1.2/1/1 | 0.9/0.9/0.9 | 0.94/0.94/1 | 1/0.94/0.94 | 1.2/1/1 | 1/1/1.2 | 1.2/1/1 |
| RsfMRI | | | | | | | | |
| Eyes | Open | No instruction | Open/ no instruction | Open | Closed | Open | Open | Open |
| Sequence | GRE-EPI | GRE-EPI | GRE-EPI | GRE-EPI | GRE-EPI | GRE-EPI | GRE-EPI | GRE-EPI |
| Phase encoding direction | A-P | P-A | A-P | A-P | P-A | P-A | P-A | P-A |
| Repetition time (ms) | 2000 | 2000 | 2000 | 2000 | 2000 | 2500 | 2500 | 2500 |
| Echo time (ms) | 31 | 30 | 30 | 30 | 30 | 30 | 30 | 30 |
| Flip angle | 80 | 75 | 90 | 90 | 90 | 80 | 80 | 80 |
| Resolution (mm, x/y/z) | 3.5/3.5/3 | 3/3/3 | 3.75/3.75/5 | 4/4/4 | 3.44/3.44/3.5 | 3.31/3.31/3.2 | 3.31/3.31/3.2 | 3.31/3.31/3.2 |
| Slice order | Interleaved | Ascending | Interleaved | Interleaved | Interleaved | Ascending | Ascending | Ascending |
| Slice gap (mm) | 0 | 1 | 0 | 0 | 0 | 0.8 | 0.8 | 0.8 |
| volumes | 150 | 210 | 150 | 180 | 150 | 240 | 244 | 240 |

**Abbreviations**

FSPGR, fast-spoiled gradient echo; GRE-EPI, gradient echo echo-planar imaging; IoPPN, Institute of Psychiatry, Psychology and Neuroscience, King's College London; JHU, Johns Hopkins University; KMU, Kanazawa Medical University; Kyoto, Kyoto University; MPRAGE, magnetization-prepared rapid gradient echo; MP2RAGE, magnetization-prepared 2 rapid acquisition gradient echoes; OSK2, Osaka University MRI2; OSK3, Osaka University MRI3; RsfMRI, resting-state functional magnetic resonance imaging; Tokyo, The University of Tokyo; TYM, University of Toyama.

**Supplemental Table S9. Thresholds of ICA-denoising at each scanner/institution.**

| Threshold | IoPPN | JHU | KMU | Kyoto | OSK2 | OSK3 | Tokyo | TYM |
|---|---|---|---|---|---|---|---|---|
| HF | 0.2 | 0.3 | 0.3 | 0.3 | 0.3 | 0.3 | 0.3 | 0.5 |
| NB | 0.6 | 0.7 | 0.7 | 0.7 | 0.7 | 0.7 | 0.7 | 0.6 |
| SL | 2 | 2 | 2 | 2 | 2 | 2 | 2 | 2 |

**Abbreviations**

ICA, independent component analysis; HF, high-frequency power ratio; IoPPN, Institute of Psychiatry, Psychology and Neuroscience, King's College London; JHU, Johns Hopkins University; KMU, Kanazawa Medical University; Kyoto, Kyoto University; NB, non-gray matter index; OSK2, Osaka University MRI2; OSK3, Osaka University MRI3; SL, slice dependency; Tokyo, The University of Tokyo; TYM, University of Toyama.

**Supplemental table S10. Matching to templates.**

| | IC number | Visual inspection | Correlation coefficient (r) |
|---|---|---|---|
| Matching to the BGN template (Supplemental Figure S3a) | | | |
| | 9 | BGN-Str | 0.65 |
| | 20 | BGN-MbThal | 0.47 |
| | 42 | CSF (ventricle) | 0.43 |
| | 69 | CSF | 0.22 |
| | 66 | White matter | 0.13 |
| Matching to the MTLN template (Supplemental Figure S3b) | | | |
| | 28 | MTLN | 0.58 |
| | 38 | Temporal pole (susceptibility artifact) | 0.22 |
| | 59 | Noise | 0.13 |
| | 46 | CSF | 0.1 |
| | 69 | CSF | 0.09 |
| Matching to the SN template (Supplemental Figure S3c) | | | |
| | 17 | SN-ACC | 0.56 |
| | 53 | SN-Ins | 0.38 |
| | 44 | MPFC | 0.24 |
| | 66 | White matter | 0.22 |
| | 21 | Bilateral supramarginal gyrus | 0.14 |

All 80 independent components (ICs) from meta-independent component analysis (ICA) were matched to the templates for each network of interest (NOI). The ICs with the top five correlation coefficients are displayed. This clearly shows that the five visually identified NOIs have the highest correlation coefficients for their templates.

Abbreviations

ACC, anterior cingulate cortex; BGN, basal ganglia network; CSF, cerebrospinal fluid; MbThal, midbrain and thalamus; MPFC, medial prefrontal cortex; MTLN, medial temporal lobe network; SN, salience network; Str, striatum.

**Supplementary Figure S1. ICA-denoising classification of all 62 components of an example participant (IC1–16)**

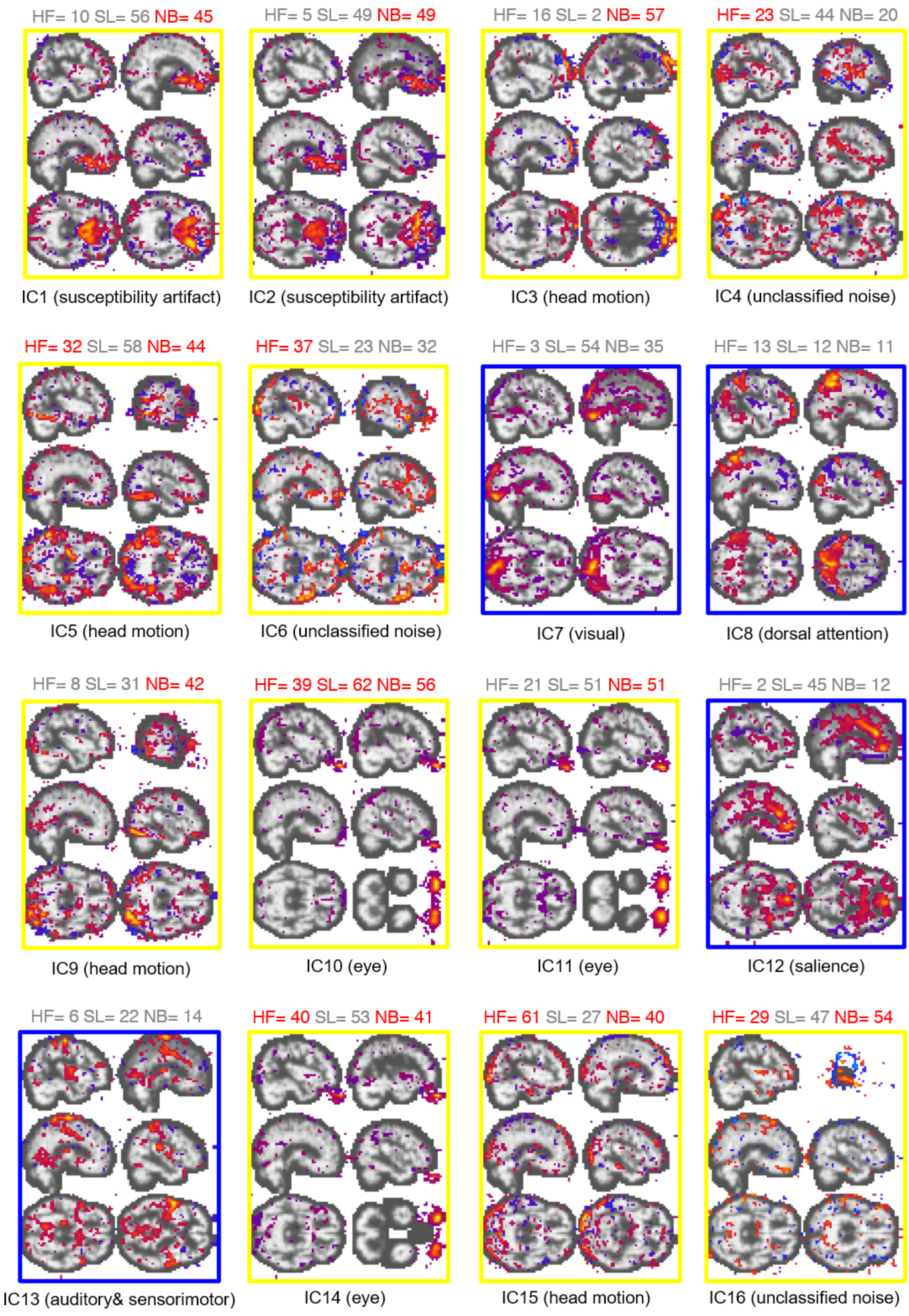

**Supplementary Figure S1. ICA-denoising classification of all 62 components of an example participant (IC17–32)**

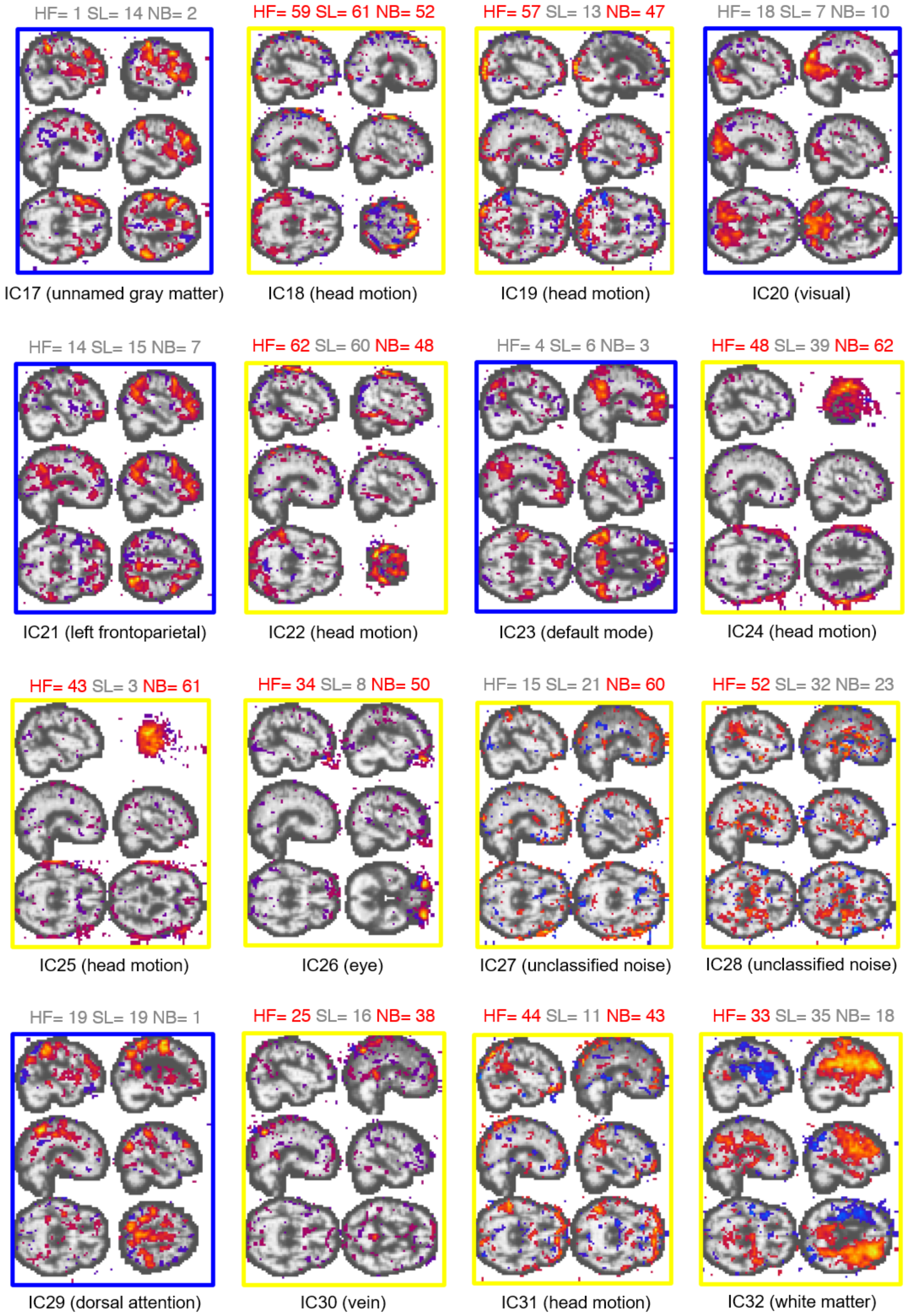

**Supplementary Figure S1. ICA-denoising classification of all 62 components of an example participant (IC33–48)**

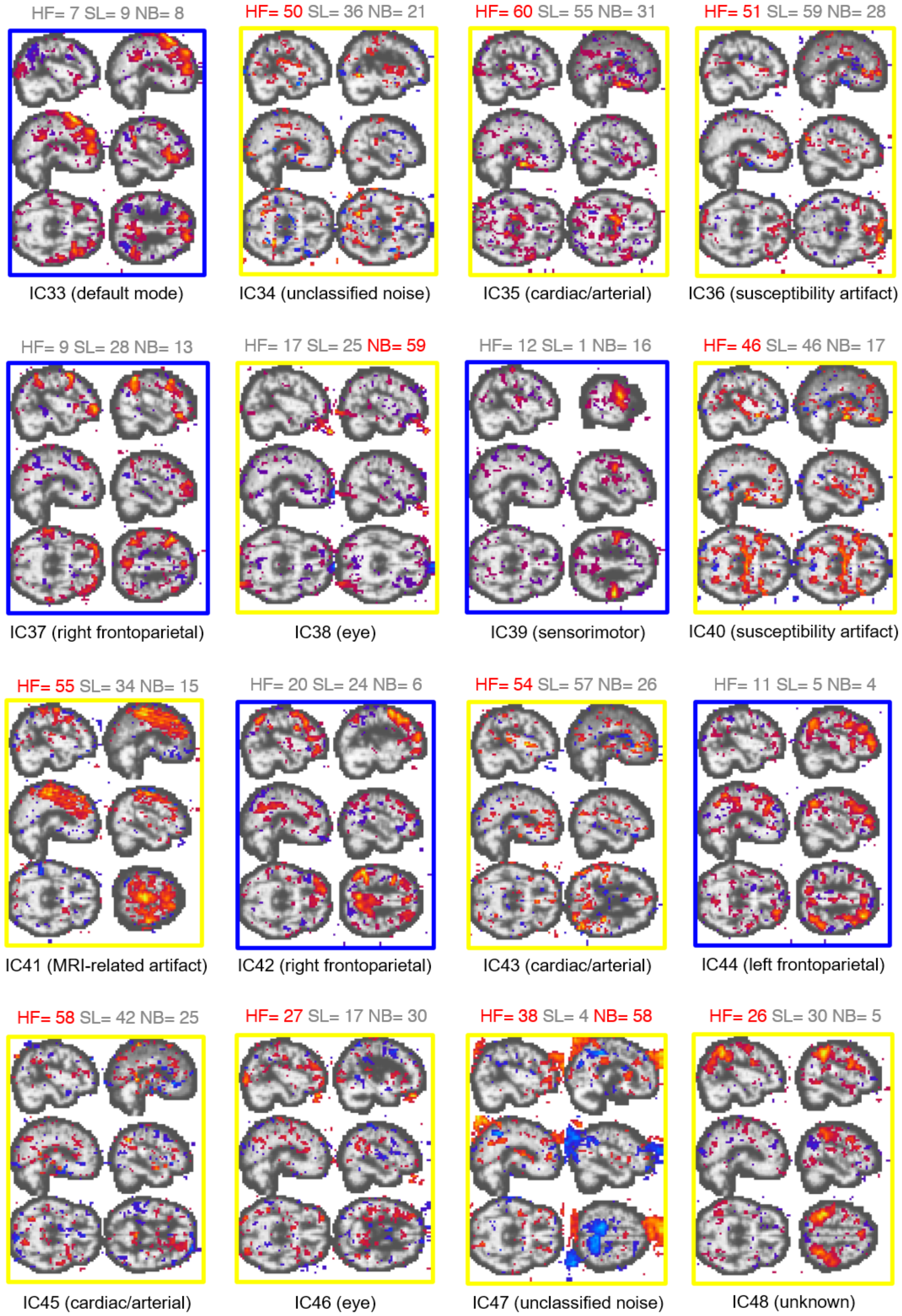

**Supplementary Figure S1. ICA-denoising classification of all 62 components of an example participant (IC49–62)**

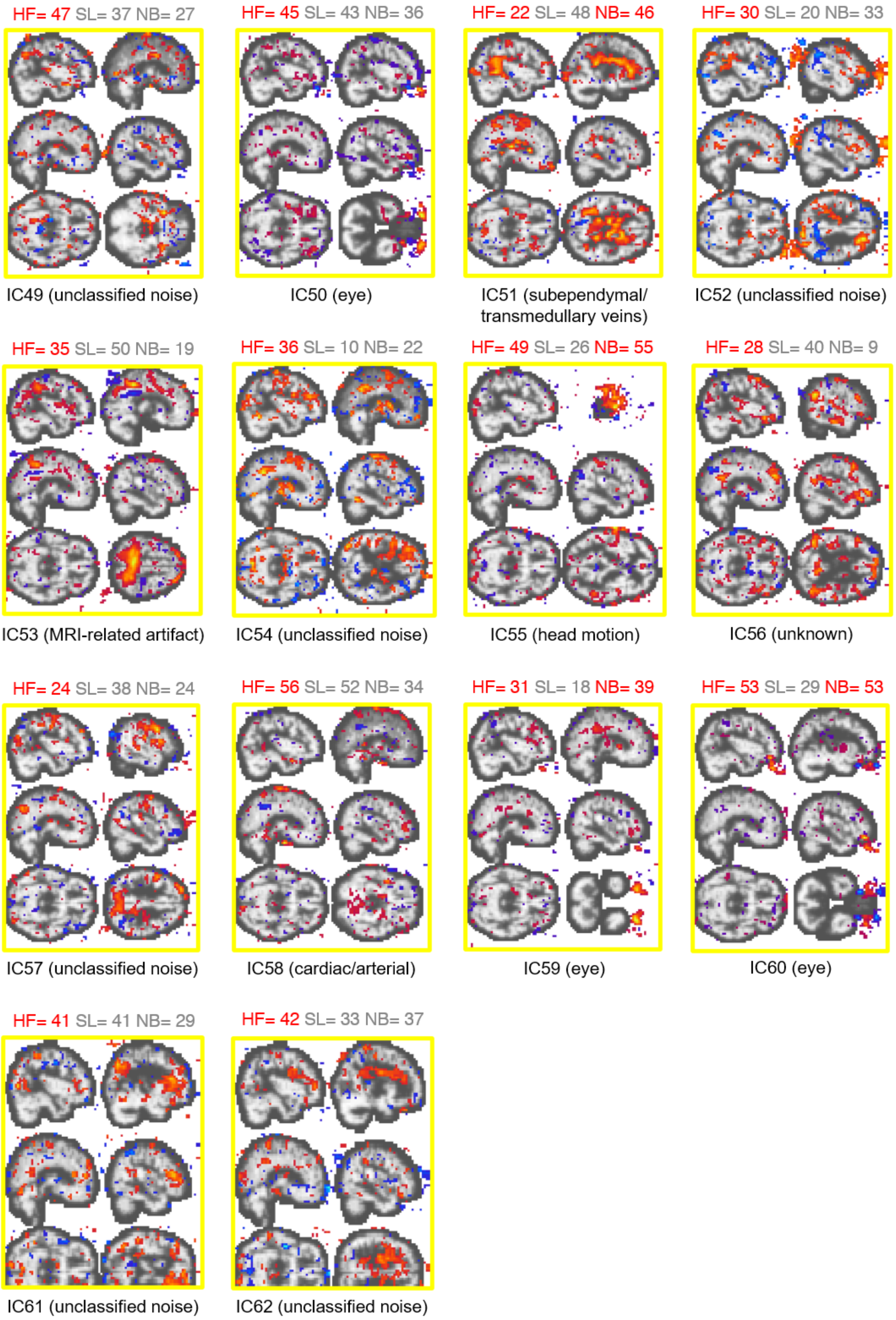


After single-subject ICA was applied to the rsfMRI data, the independent components (ICs) were classified as noise based on the following criteria:

1) high-frequency power ratio (>0.1 Hz up to 0.2 Hz or the Nyquist frequency for the repetition time [TR]) relative to <0.1 Hz (HF)

2) non-gray matter involvement index (NB)

3) slice dependency index calculated as the ratio of within- and between-slice spatial high-frequency components (SL)

The ICs surrounded by blue frames are neuronal components and the ICs with yellow frames are noise components. The numbers indicate the ranks among all 62 ICs, with smaller ranks indicating higher quality. The red ranks indicate noise classification.

Abbreviations

ICA, independent component analysis; rsfMRI, resting-state functional magnetic resonance imaging.

**Supplementary Figure S2. ICA-denoising removed low frequency noises.**

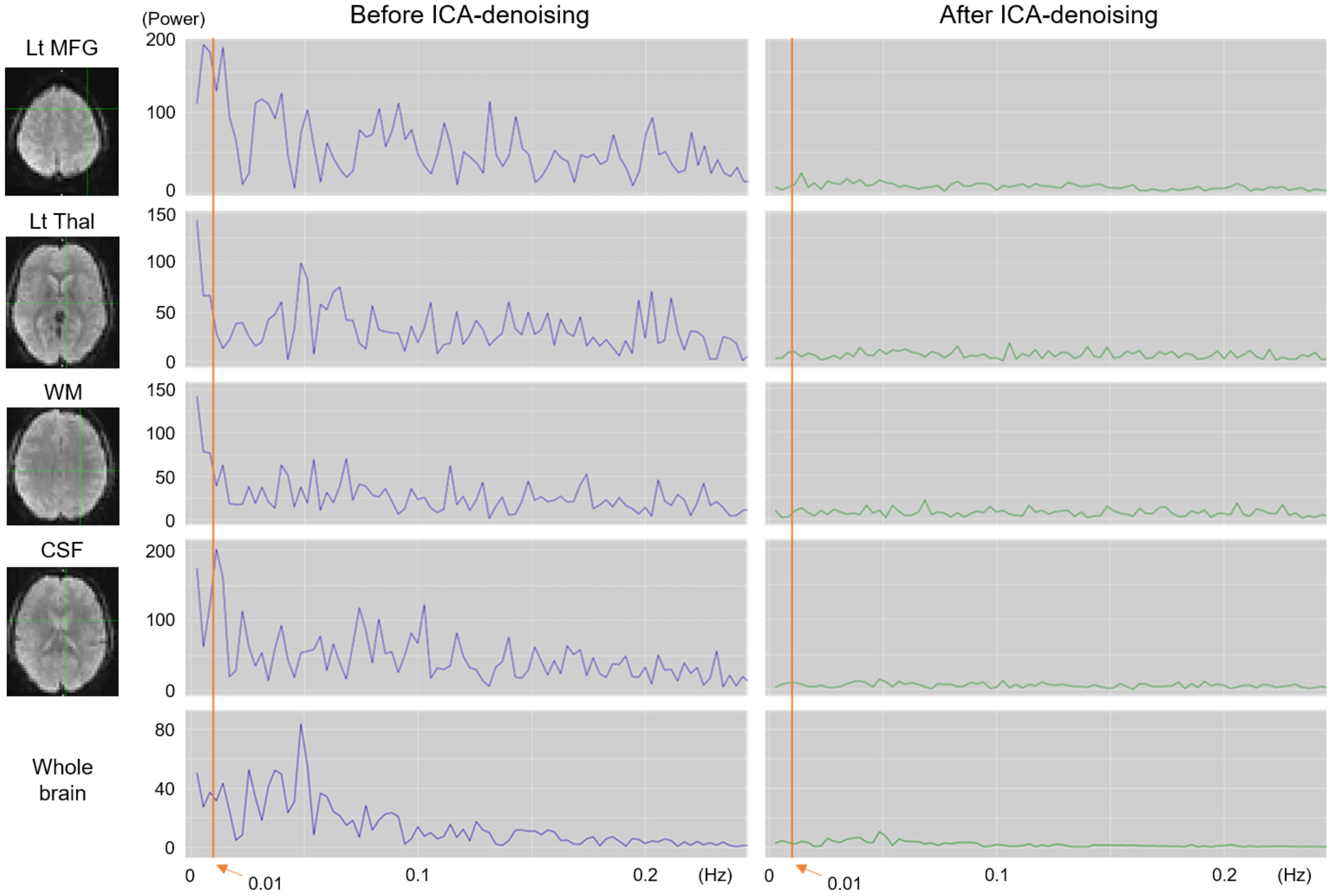

Power spectra analysis of voxelwise (1st to 4th row) and whole brain time courses revealed that low frequency noises below 0.01Hz (orange line) were efficiently removed by ICA-denoising.

**Abbreviations**

CSF, cerebrospinal fruid (lateral ventricle); ICA, independent component analysis; Lt, left; MFG, middle frontal gyrus; Thal, thalamus; WM, white matter (semioval center);

**Supplemental Figure S3. Templates for each NOI.**

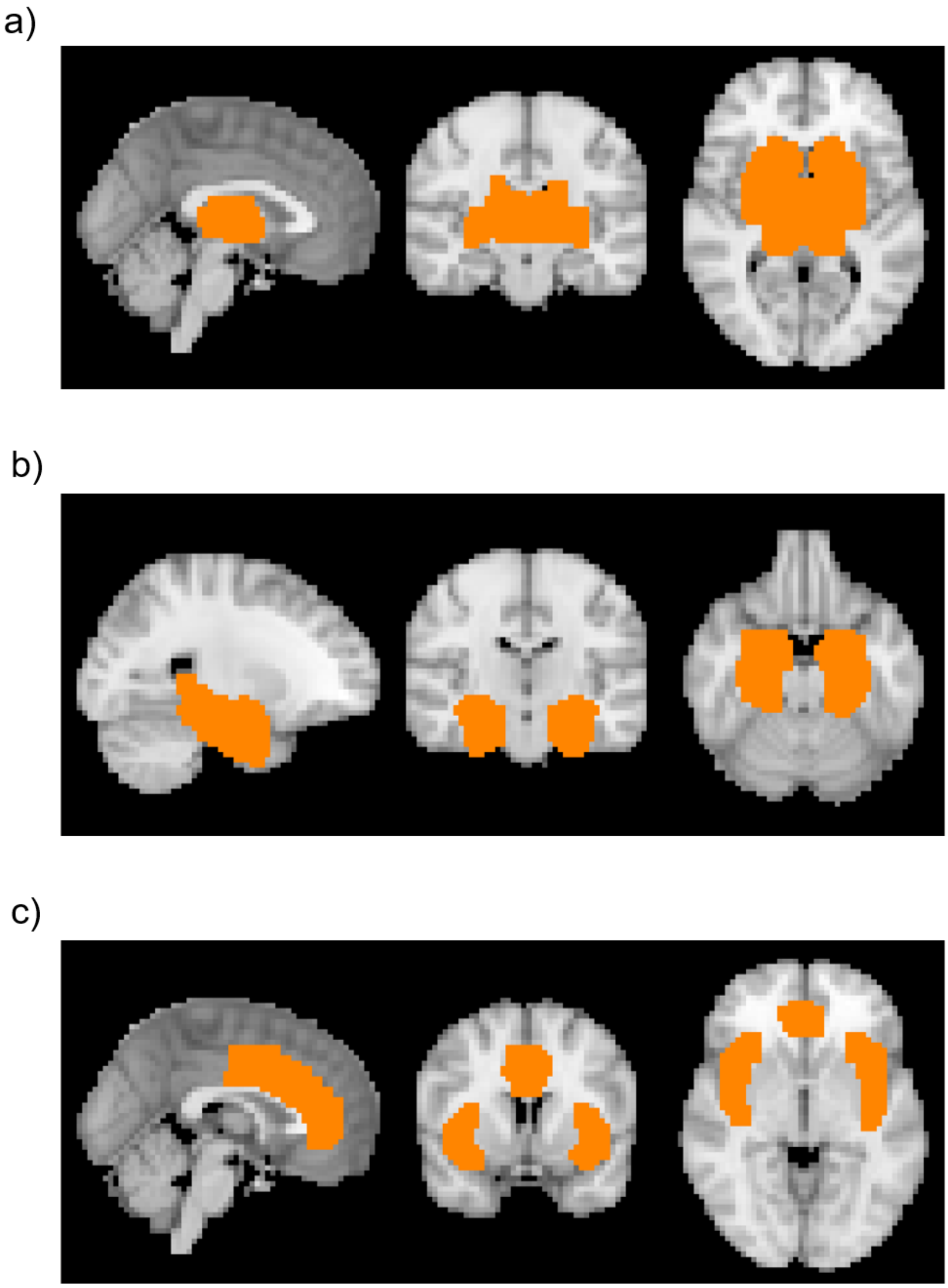


The templates for each network of interest (NOI) are shown. a) The template for the basal ganglia network (BGN) was created by combining the bilateral thalamus, caudate, putamen, and accumbens from the Harvard-Oxford Subcortical Structural Atlas. b) The salience network (SN) template was created by combining the anterior cingulate cortex (ACC) and bilateral insula from the atlas. c) The medial temporal lobe network (MTLN) template was created by combining the bilateral amygdala, hippocampus, and anterior/posterior parahippocampal gyri.

**Supplemental Figure S4. Harmonization by ComBat effectively removed site/scanner effects.**

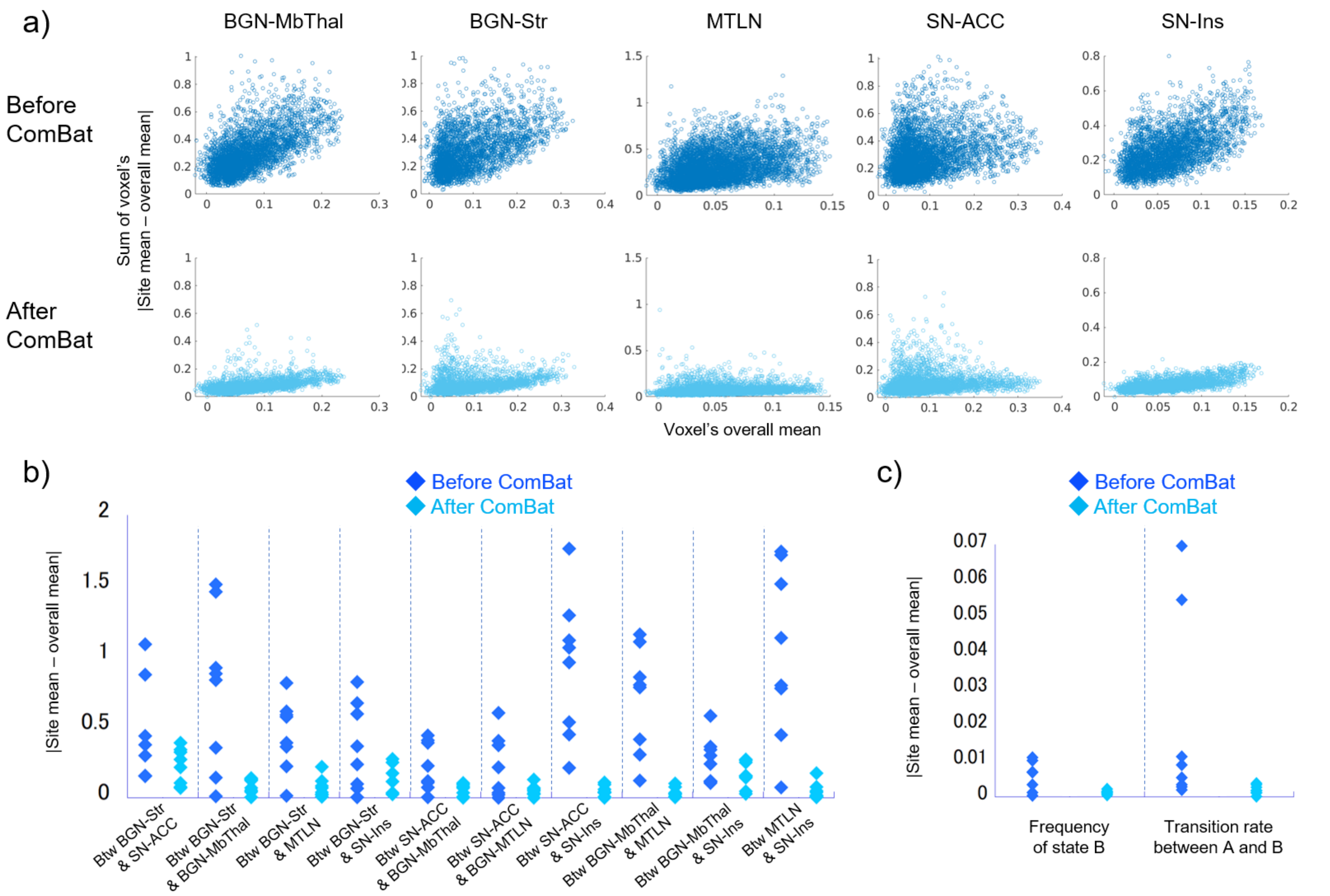

**a) Effect of ComBat on the spatial maps of the within-network connectivity analysis.**

The sum of each voxel's |site mean − overall mean| was plotted against the overall mean of each voxel, controlling for age, sex, and tSNR.

**b) Effect of ComBat on the Z-transformed partial correlation coefficients of the between-network connectivity analysis.**

The |site mean − overall mean| of the correlation coefficients was plotted for each NOI-pair, controlling for age, sex, and tSNR.

**c) Effect of ComBat on the frequency of state B and transition rate between states A and B of the energy landscape analysis.**

The |site mean − overall mean| of each index was plotted controlling for age, sex, and tSNR.

**Abbreviations**

ACC, anterior cingulate cortex; BGN, basal ganglia network; Btw, between; Ins, insula; MbThal, midbrain and thalamus; MTLN, medial temporal lobe network; SN, salience network; Str, striatum; tSNR, temporal signal-to-noise ratio.

**Supplemental Figure S5. Reduced within-network connectivity in patients with UHR, FEP, and ChrP at liberal thresholds.**

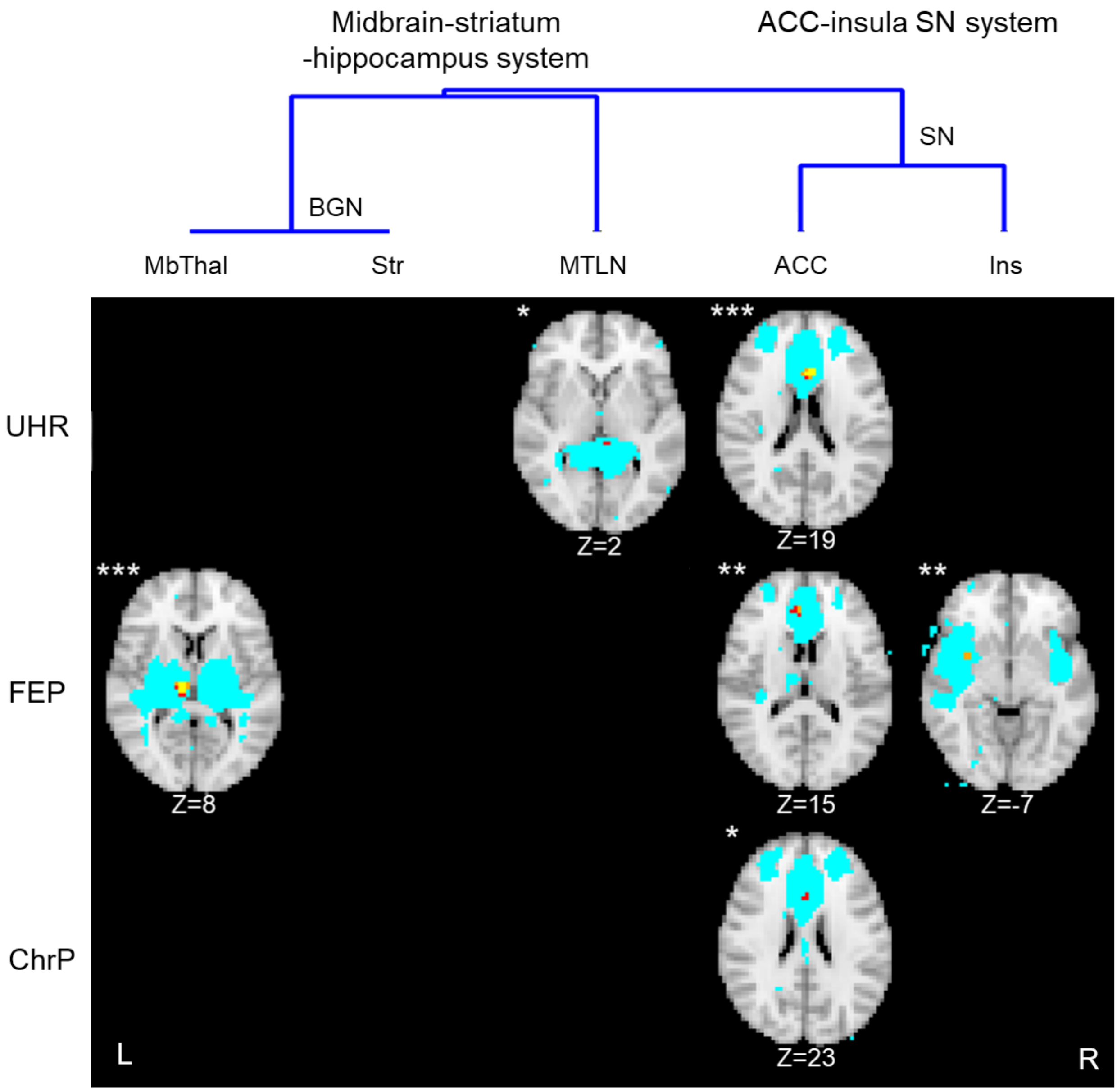


Patients with UHR (n=29) were compared to matched HC (n=25), FEP (n=81) to matched HC (n=109), and ChrP (n=99) to matched HC (n=145). Yellow and *** indicate a significant reduction of connectivity in patients at $p < 0.05$, family-wise error (FWE) corrected for voxels, contrasts, and networks. Orange and ** indicate trend-level significance at $p < 0.1$, FWE for voxels, contrasts, and networks. Red and * indicate significance at $p < 0.05$, FWE for voxels and contrasts. Light blue indicates the NOIs.

**Abbreviations**

ACC, anterior cingulate cortex; BGN, basal ganglia network; ChrP, chronic schizophrenia; FEP, first-episode psychosis; HC, healthy controls; Ins, insula; MbThal, midbrain and thalamus; MTLN, medial temporal lobe network; NOI, network of interest; SN, salience network; Str, striatum; UHR, ultra-high-risk.

**Supplemental Figure S6. Correlations between positive symptom severity and within- and between-network connectivity at liberal thresholds.**

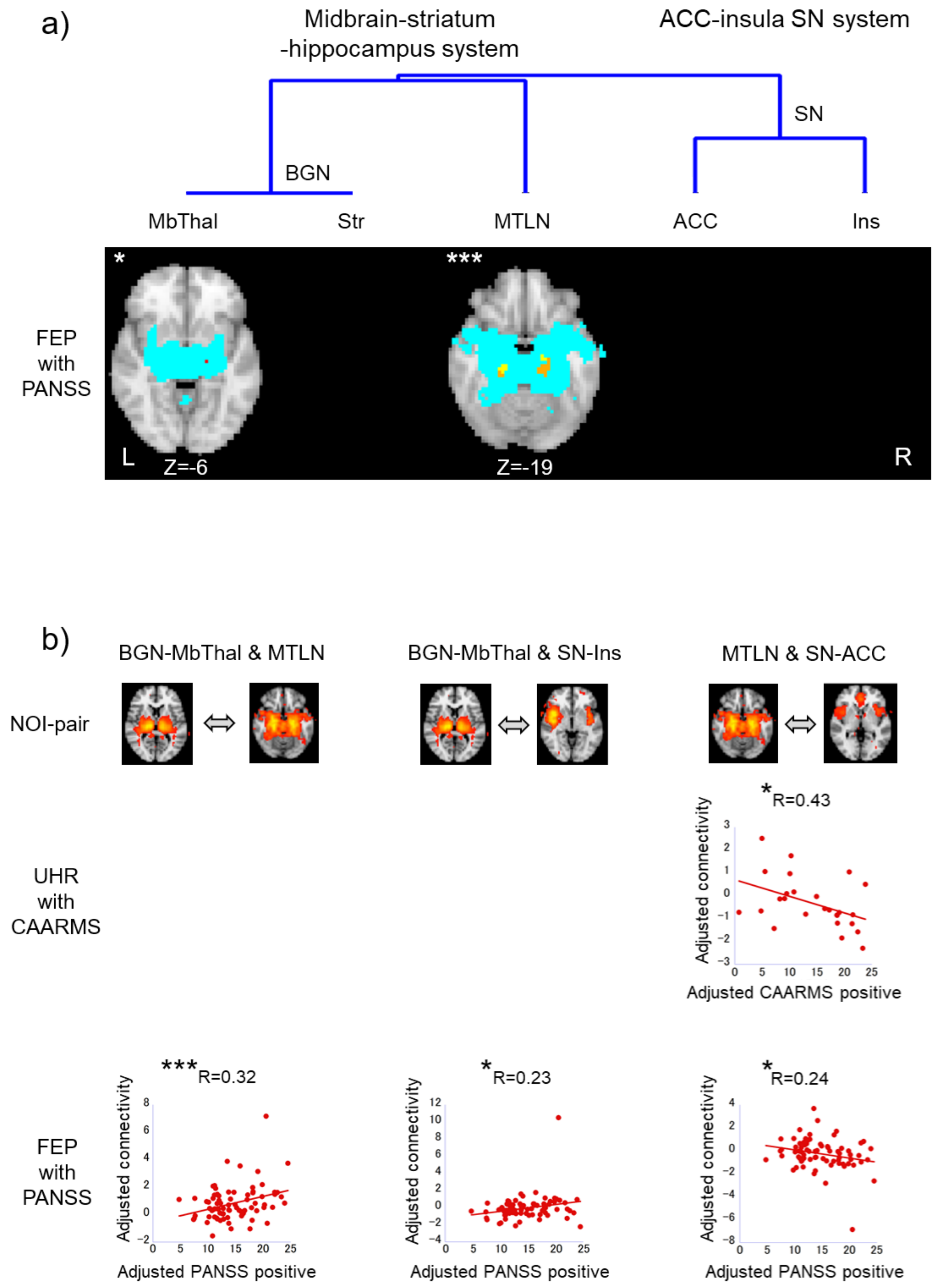

**a) Positive correlations between the PANSS positive scale and within-network connectivity in FEP (n=76)**

Yellow and *** indicate significance at $p < 0.05$, family-wise error (FWE) corrected for voxels, contrasts, and networks. Orange indicates trend-level significance at $p < 0.1$, FWE for voxels, contrasts, and networks. Red and * indicate significance at $p < 0.05$, FWE for voxels and contrasts. Light blue indicates the NOIs.

**b) Correlations between the positive symptom scales and between-network connectivity in UHR (n=26) and FEP (n=76).**

*** significance at $p < 0.05$, FWE for contrasts and network-pairs. * significance at $p < 0.05$, FWE for contrasts. CAARMS and PANSS scores and connectivity values were adjusted for age, sex, and tSNR.

**Abbreviations**

ACC, anterior cingulate cortex; BGN, basal ganglia network; CAARMS, Comprehensive Assessment of At-Risk Mental States; FEP, first-episode psychosis; Ins, insula; MbThal, midbrain and thalamus; MTLN, medial temporal lobe network; NOI, network of interest; PANSS, Positive and Negative Syndrome Scale; SN, salience network; Str, striatum; tSNR, temporal signal-to-noise ratio; UHR, ultra-high-risk.

**Supplemental Figure S7. Effect of comorbidity on correlational analysis**

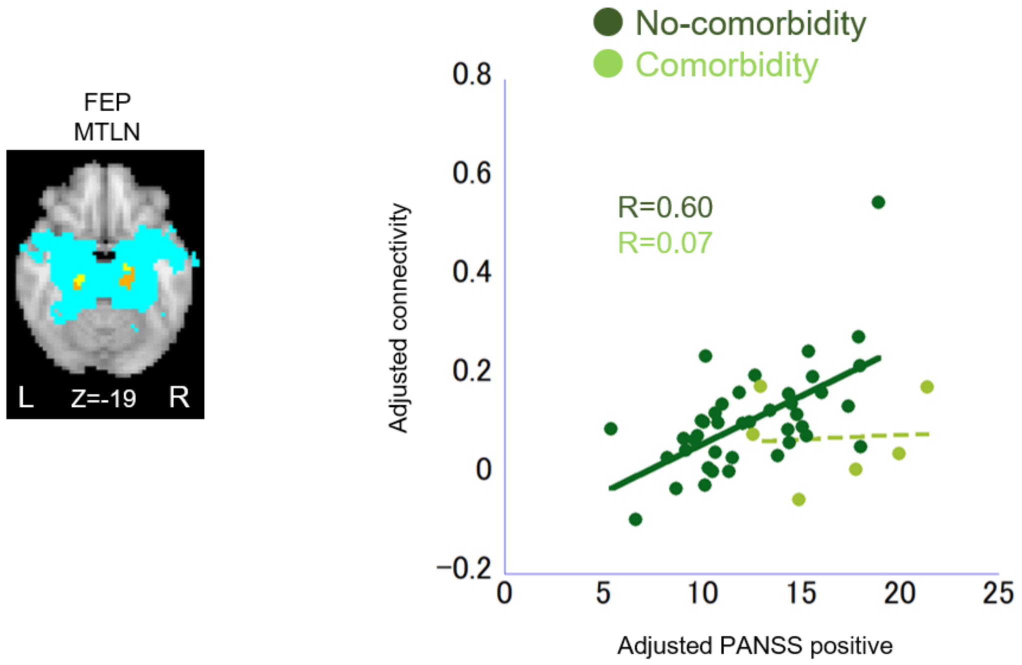


The regression coefficient between the PANSS positive scale and the within-network connectivity of the MTLN was larger at the trend-level in patients without (n=41) than in those with (n=6) comorbidities ($p = 0.08$, family-wise error corrected for contrasts). PANSS scores and connectivity values were adjusted for age, sex, and tSNR.

**Abbreviations**

FEP, first-episode psychosis; MTLN, medial temporal lobe network; PANSS, Positive and Negative Syndrome Scale; tSNR, temporal signal-to-noise ratio.

**Supplemental Figure S8. Effects of smoking on the correlational analyses**

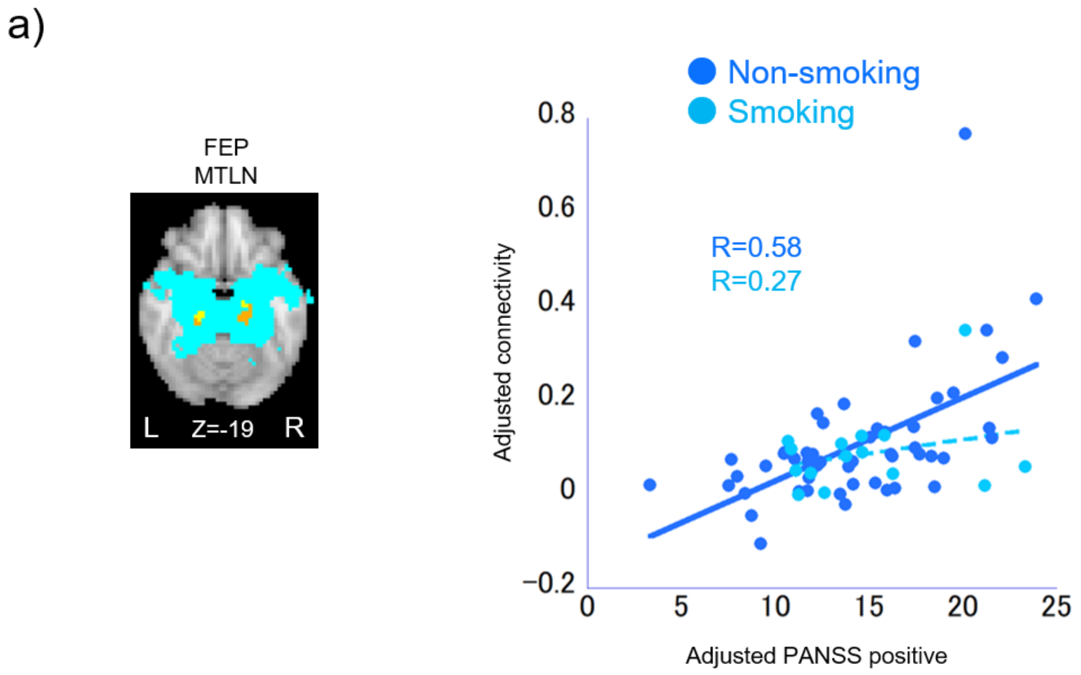


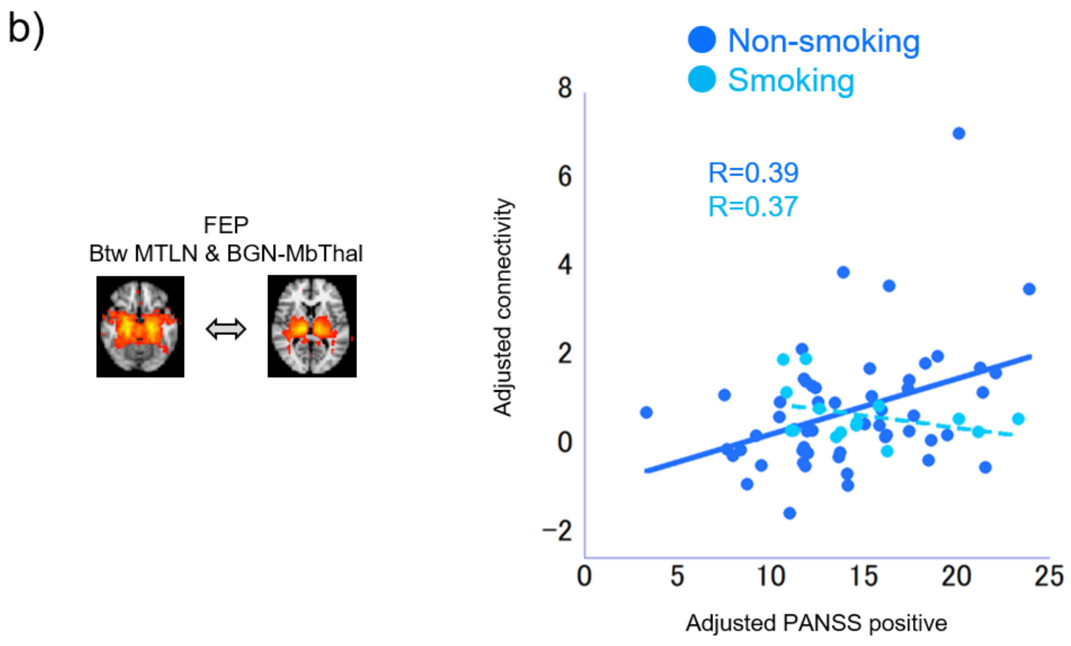


a) The regression coefficient between PANSS positive scale and the within-network connectivity of the MTLN was larger at the trend-level in non-smokers (n=54) than in smokers (n=15) (p = 0.07, family-wise error [FWE] corrected for contrasts). PANSS scores and connectivity values were adjusted for age, sex, and tSNR.

b) The regression coefficient between PANSS and the connectivity between the MTLN and BGN-MbThal was larger at the trend level in non-smokers than in smokers ($p = 0.05$, FWE for contrasts).

**Abbreviations**

BGN-MbThal, midbrain-thalamus part of the basal ganglia network; FEP, first-episode psychosis; MTLN, medial temporal lobe network; PANSS, Positive and Negative Syndrome Scale; tSNR, temporal signal-to-noise ratio.
